# Distinct Cervicovaginal Cytokine Signatures Associated with Reproductive Tract Infections and Vaginal Dysbiosis Across Diverse Settings

**DOI:** 10.64898/2026.06.26.26356651

**Authors:** Micaela Lurie, Tania Crucitti, Musalula Sinkala, Ramla F. Tanko, Aina Harimanana, Katherine Gill, Linda-Gail Bekker, Janneke HHM van de Wijgert, Bich-Tram Huynh, Camille Fortas, Stéphanie Ramboarina, Theodora Mayouya Gamana, Rindra Randremanana, Reziky T. Mangahasimbola, Soamiangaly Randrianjatovo, Noel H. Ratovonirina, Chido Dziva Chikwari, Tinashe Mwaturura, Katharina Kranzer, Nicola Thomas, Anda Madikida, Karabo Mahlangu, David Anderson, Emma Harding-Esch, Constance Macworth-Young, Edina Sinanovic, Elise Smith, Ayako Honda, Fezile Kumalo, Monalisa T. Manhanzva, Tanya Pidwell, Jo-Ann S. Passmore, Lindi Masson, the GIFT study group

**Author notes:** Co-last authors (Jo-Ann S. Passmore, Lindi Masson). Corresponding author: Prof Jo-Ann Passmore, Centre for Epidemic Response and Innovation (CERI), School of Data Science and Computational Thinking, Stellenbosch University, Stellenbosch, 7600.

## Abstract

**Background:** Reproductive tract infections (RTIs) and bacterial vaginosis (BV) cause significant reproductive morbidity but often remain undetected. We evaluated cervicovaginal cytokine signatures associated with RTIs and vaginal dysbiosis in women from South Africa, Madagascar, and Zimbabwe.

**Methods:** Vaginal swabs from 676 sexually-active women (18–35 years) were tested for *Chlamydia trachomatis* (CT), *Neisseria gonorrhoeae* (NG), *Trichomonas vaginalis* (TV), *Mycoplasma genitalium* (MG), *Candida* spp., and BV. IL-1α, IL-1β, and IP-10 were measured by ELISA; associations were assessed using multivariable regression and population attributable fractions.

**Results:** BV prevalence was 50.4% (337/668), followed by CT (15.0%), TV (11.8%), NG (5.7%), MG (4.5%), and *Candida* spp. (6.8%). BV dominated elevated IL-1α/IL-1β, accounting for >60% of high cytokine responses. Intermediate microbiota (Nugent 4–6) showed similar inflammatory profiles and, with BV, reduced IP-10. Independently, NG was associated with elevated IL-1α/IL-1β; CT with elevated IL-1β/IP-10; TV with elevated IP-10; *Candida* spp. with all cytokines; MG showed no association. Most infections were asymptomatic, eliciting similar inflammatory profiles to symptomatic cases. Pathogen-specific inflammatory signatures were consistent across countries.

**Conclusions:** RTIs and dysbiosis elicited consistent inflammatory signatures across countries. Their high prevalence in asymptomatic women highlights the potential of host-response biomarkers to identify undetected genital inflammation (ClinicalTrials.gov: NCT05723484).

**Lay summary:** Reproductive tract infections and bacterial vaginosis (BV; a common condition in which the normal protective bacteria in the vagina are replaced by a mix of other bacteria) are major causes of poor reproductive health, but many women have no symptoms and remain undiagnosed. In this study, we measured inflammatory proteins in vaginal samples from 676 women in South Africa, Madagascar, and Zimbabwe and examined how these markers were associated with infections and changes in the vaginal microbiota. We found that BV was the strongest driver of genital inflammation and that women with intermediate vaginal microbiota, often considered a transitional microbial state, showed similar inflammatory profiles. Different infections were associated with distinct inflammatory patterns, but many women with substantial inflammation had no symptoms. Importantly, these inflammatory responses were broadly similar across all three countries. Together, these findings highlight important limitations of symptom-based diagnosis and support the development of host-response diagnostics that can identify by reproductive tract infections and vaginal dysbiosis, including in women who would otherwise remain undiagnosed.

## Introduction

Reproductive tract infections (RTIs), including sexually transmitted infections (STIs), vulvovaginal candidiasis (VVC), and bacterial vaginosis (BV), cause significant reproductive morbidity and mucosal inflammation (1). Besides causing pelvic inflammatory disease, infertility, and adverse pregnancy outcomes, RTIs increase HIV susceptibility by promoting mucosal immune activation and disrupting epithelial barrier integrity (2–5). Key causative pathogens, including *Chlamydia trachomatis* (CT), *Neisseria gonorrhoeae* (NG), *Trichomonas vaginalis* (TV), *Mycoplasma genitalium* (MG), *Candida* spp., and BV, are highly prevalent in sub-Saharan Africa (6–8). BV, characterized by a shift from protective Lactobacillus-dominated microbiota to diverse anaerobes, is highly prevalent globally (9, 10). Candida spp. can exist as commensals or cause symptomatic VVC (11), inducing IL-1 cytokine secretion (12–14). This highlights the importance of evaluating these multi-kingdom conditions within a unified framework of mucosal inflammation.

Despite this high burden, many infections are asymptomatic and missed by syndromic management (1). While BV, TV, and VVC often present with discharge, CT and NG are frequently asymptomatic (15). Yet, both vaginal and cervical RTIs induce inflammatory cytokine responses poorly reflected by symptoms (16). This dissociation highlights the need to define host inflammatory profiles associated with distinct RTIs across diverse settings.

In East and Southern African cohorts, STIs and BV consistently elevate cervicovaginal IL-1α and IL-1β (17, 18). Conversely, BV suppresses the chemokine IP-10, creating a distinct inflammatory signature (17–20). These biomarker signatures are detectable in asymptomatic women (16), highlighting their utility in inflammation-based screening to identify undetected infections and dysbiosis.

Since RTI prevalence (6), vaginal microbiota composition (21), and host immunity vary across populations (22, 23), it is important to determine whether these inflammatory signatures are consistent across diverse settings. Here, we evaluated cervicovaginal cytokine profiles in relation to laboratory-confirmed RTIs, considering both symptomatic and asymptomatic cases. Focusing on IL-1α, IL-1β, and IP-10 (17, 19), we aimed to identify the RTIs most strongly associated with inflammation, extend analyses to include Candida spp., and assess the consistency of these signatures across diverse African settings.

## Methods and Materials

### Study population, enrolment and ethics

We enrolled 676 non-pregnant, sexually-active women aged 18–35 years from South Africa (n=225), Zimbabwe (n=226), and Madagascar (n=225) (Supplementary Figure 1; NCT05723484) (24). Participants were recruited at the Desmond Tutu Health Foundation Masiphumelele clinical research site in Cape Town, South Africa; the Spilhaus Family Planning Centre in Harare, Zimbabwe; and the Centre Hospitalier Universitaire de Gynécologie et d’Obstétrique Gynéco-Obstétricaux de Befelatanana in Antanarivo, Madagascar. In brief, eligible participants were non-pregnant, sexually-active women aged 18–35 years who were willing to provide written informed consent and undergo pelvic examination. Exclusion criteria included active menstruation, vaginal bleeding, pregnancy (confirmed by urine pregnancy test at screening), having given birth or had a miscarriage in the past 6 weeks, or having used any vaginal insert or douche in the previous 48 hours. Blood was collected for HIV testing according to national algorithms. Women living with HIV were referred for antiretroviral therapy according to national guidelines but were not excluded.

### Clinical evaluation and specimen collection

At enrolment, participants completed structured interviews and underwent general and gynaecological examinations (24). Seven lateral vaginal wall swabs were collected for RTI testing and cytokine analysis. Participants were managed syndromically according to WHO-adapted national guidelines, with treatment and partner notification provided on-site. Socio-demographic, behavioural, and clinical data (including sexual behavior and contraceptive use) were captured using REDCap (24).

### RTI diagnosis

Vaginal swabs were tested for discharge-causing RTIs centrally at the Institut Pasteur de Madagascar (IPM). CT and NG were detected using the Presto CT/NG qPCR platform (Goffin Molecular Technologies), while TV, MG, and semen (Y-chromosome) (26) were detected using validated in-house qPCR assays (24). BV was diagnosed by Nugent scoring of Gram-stained smears (9). Detection of *Candida* spp. utilized an in-house multiplex qPCR assay targeting the ITS2 ribosomal DNA region to detect *C. albicans*, *C. parapsilosis*, and *C. guilliermondii* (24).

### Cytokine measurements

Lateral vaginal swabs stored at −80 °C were thawed and eluted in 1 ml phosphate-buffered saline (PBS). Eluates were assayed for IL-1α, IL-1β, and IP-10 using Quantikine ELISA kits (R&D Systems, USA) (17), according to manufacturer instructions. Five inter-plate vaginal samples with known cytokine concentrations were included on each plate, and inter-plate coefficients of variation ≤ 15% were considered acceptable. The lower and upper limits of detection were 3.9-250 pg/ml for both IL-1α and IL-1β, and 7.8-500 pg/ml for IP-10. All eluates were analysed neat. Standards were run in duplicate while vaginal eluates from each woman were run in a single well.

## Statistical Analysis

Women were classified as symptomatic if they reported abnormal vaginal discharge or lower abdominal pain (1). For descriptive analyses, RTI status was categorized as: uninfected controls (negative for all STIs/*Candida*; Nugent 0–3), STI-only (STI-positive, Nugent 0–3, *Candida*-negative), BV-only (Nugent 7–10, STI/*Candida*-negative), intermediate microbiota only (Nugent 4–6, STI/*Candida*-negative), and *Candida*-only (*Candida*-positive, STI-negative, Nugent 0–3). Cytokines were compared using Mann-Whitney U or Kruskal-Wallis tests.

Poisson regression with robust variance was used to estimate relative risks (RRs) and 95% confidence intervals (CIs) for elevated cytokines (defined as the upper quartile of log10-transformed distributions). Separate models were built for each cytokine, using women without the condition as reference and adjusting for co-infections. Geographic variation was evaluated using multi-level mixed-effects regression models with interaction terms for country, adjusting for co-infections, vaginal hygiene, HIV status, contraceptive use, and Y-chromosome detection.

Population attributable fractions (PAFs) were calculated using the formula Pi(RRi−1)/[1+Pi(RRi−1)], where Pi represents the prevalence of each exposure and RRi represents the relative risk of elevated cytokine concentrations associated with that exposure. Statistical significance was defined as p<0.05. All analyses were performed using R, GraphPad Prism v10.4.0, and MATLAB 2024b.

## Ethics

Ethical approval for this study was obtained from the University of Cape Town Human Research Ethics Committee (UCT HREC; reference numbers 366/2022 and 799/2021), the Medical Research Council of Zimbabwe (MRCZ; approval number A/2966), the Comité d’Éthique pour la Recherche Biomédicale (CERBM), Madagascar, and the London School of Hygiene & Tropical Medicine ethics committee (LSHTM reference 28046). All participants provided written informed consent.

## Results

Of 676 enrolled women, laboratory-confirmed STI/*Candida* results were available for 672, and BV status for 669 (Supplementary Figure 1). South African participants were younger (median 24.9 years) than those in Madagascar (26.5 years) and Zimbabwe (28.6 years) (Table 1; p<0.0001). HIV prevalence was 20.0% (45/225) in South Africa, <u>0.4%</u> (1/225) in Madagascar, and 7.5% (17/226) in Zimbabwe (Table 1; p<0.0001). BV (Nugent 7–10) was the most prevalent condition (50.4% overall; 337/668), peaking in South Africa (57.7%; p=0.019). Most cases were asymptomatic (Supplementary Figure 2), with South Africa having a higher asymptomatic proportion than Madagascar and Zimbabwe (p<0.0001).

**Table 1.**
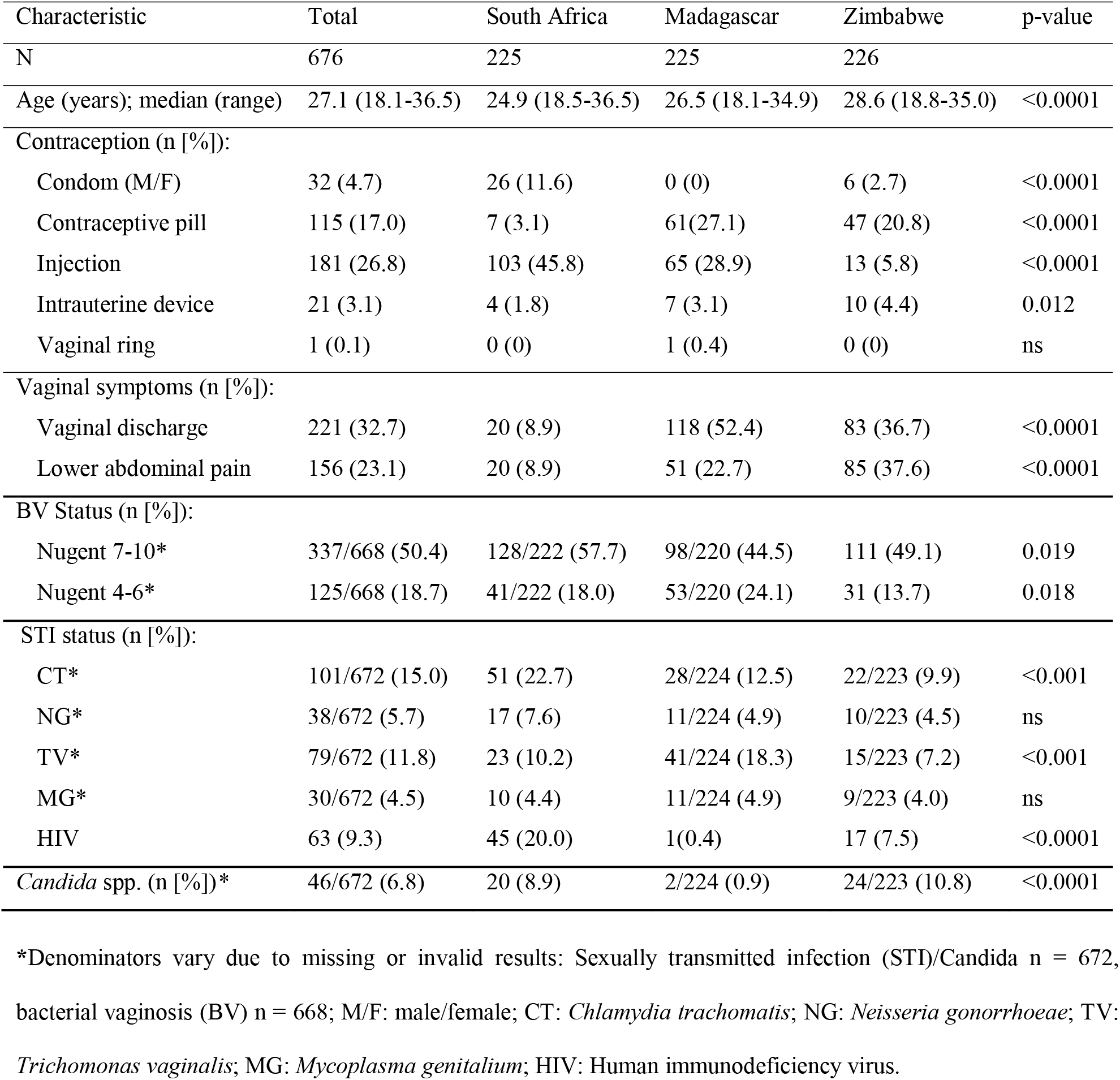
Demographic, behavioural and clinical characteristics of Genital InFlammation Test (GIFT) study participants stratified by study site.

| Characteristic | Total | South Africa | Madagascar | Zimbabwe | p-value |
| --- | --- | --- | --- | --- | --- |
| N | 676 | 225 | 225 | 226 |  |
| Age (years); median (range) | 27.1 (18.1-36.5) | 24.9 (18.5-36.5) | 26.5 (18.1-34.9) | 28.6 (18.8-35.0) | <0.0001 |
| Contraception (n [%]): |  |  |  |  |  |
| Condom (M/F) | 32 (4.7) | 26 (11.6) | 0 (0) | 6 (2.7) | <0.0001 |
| Contraceptive pill | 115 (17.0) | 7 (3.1) | 61(27.1) | 47 (20.8) | <0.0001 |
| Injection | 181 (26.8) | 103 (45.8) | 65 (28.9) | 13 (5.8) | <0.0001 |
| Intrauterine device | 21 (3.1) | 4 (1.8) | 7 (3.1) | 10 (4.4) | 0.012 |
| Vaginal ring | 1 (0.1) | 0 (0) | 1 (0.4) | 0 (0) | ns |
| Vaginal symptoms (n [%]): |  |  |  |  |  |
| Vaginal discharge | 221 (32.7) | 20 (8.9) | 118 (52.4) | 83 (36.7) | <0.0001 |
| Lower abdominal pain | 156 (23.1) | 20 (8.9) | 51 (22.7) | 85 (37.6) | <0.0001 |
| BV Status (n [%]): |  |  |  |  |  |
| Nugent 7-10* | 337/668 (50.4) | 128/222 (57.7) | 98/220 (44.5) | 111 (49.1) | 0.019 |
| Nugent 4-6* | 125/668 (18.7) | 41/222 (18.0) | 53/220 (24.1) | 31 (13.7) | 0.018 |
| STI status (n [%]): |  |  |  |  |  |
| CT* | 101/672 (15.0) | 51 (22.7) | 28/224 (12.5) | 22/223 (9.9) | <0.001 |
| NG* | 38/672 (5.7) | 17 (7.6) | 11/224 (4.9) | 10/223 (4.5) | ns |
| TV* | 79/672 (11.8) | 23 (10.2) | 41/224 (18.3) | 15/223 (7.2) | <0.001 |
| MG* | 30/672 (4.5) | 10 (4.4) | 11/224 (4.9) | 9/223 (4.0) | ns |
| HIV | 63 (9.3) | 45 (20.0) | 1(0.4) | 17 (7.5) | <0.0001 |
| *Candida* spp. (n [%])* | 46/672 (6.8) | 20 (8.9) | 2/224 (0.9) | 24/223 (10.8) | <0.0001 |

\*Denominators vary due to missing or invalid results: Sexually transmitted infection (STI)/*Candida* n = 672, bacterial vaginosis (BV) n = 668; M/F: male/female; CT: *Chlamydia trachomatis*; NG: *Neisseria gonorrhoeae*; TV: *Trichomonas vaginalis*; MG: *Mycoplasma genitalium*; HIV: Human immunodeficiency virus.

CT was the most prevalent discharge-causing STI (15.0%), followed by TV (11.8%), NG (5.7%), and MG (4.5%). Prevalence varied geographically: CT/NG peaked in South Africa, TV/MG in Madagascar, and Candida spp. in Zimbabwe. Most TV (86.1%), NG (78.4%), CT (77.1%), and MG (70.0%) cases occurred concurrently with BV (Supplementary Figure 3). Multiple STIs were rare.

### Association between RTIs and cervicovaginal cytokines

Unadjusted analyses showed that CT, NG, TV, MG, Candida spp., intermediate microbiota, and BV were associated with significantly elevated IL-1α and IL-1β compared to controls (p<0.0001; Figure 1A). TV, BV, and *Candida* spp. showed the largest effect sizes for IL-1α, while TV and BV drove the largest IL-1β increases; MG showed the smallest effects for both interleukins (Figure 1B).

**Figure 1.**
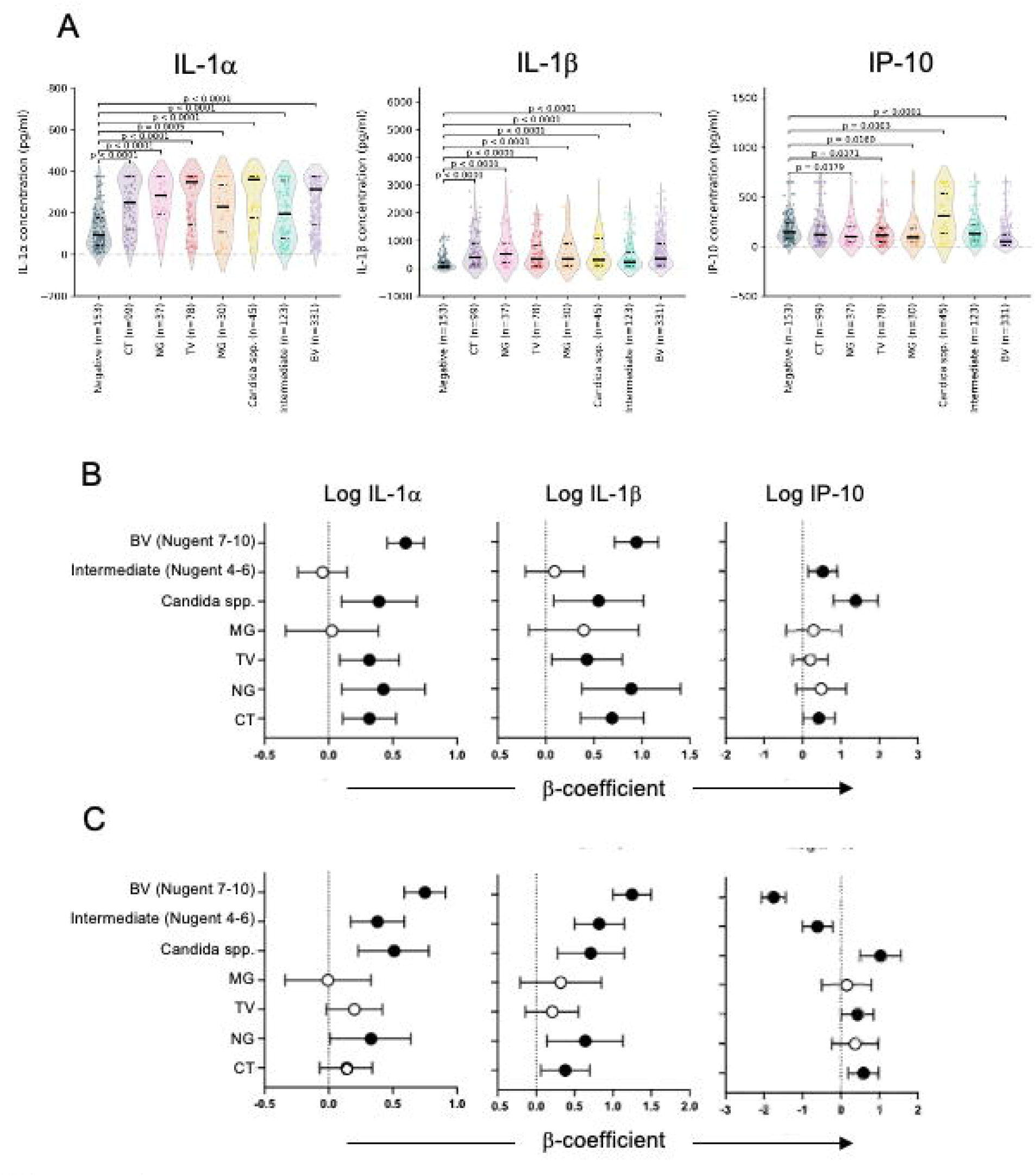
Cytokine distributions and effect-size estimates for genital infections. Cytokine concentrations were log10-transformed prior to analysis. (A) Violin plots of IL-1α, IL-1β and IP-10 by infection group. Each violin illustrates the full distribution of individual values, with the median (solid line) and interquartile range (dashed lines) overlaid. Colours indicate infection status: Negative (dark blue), CT (purple), NG (pink), TV (red), MG (orange), *Candida* spp. (yellow), intermediate microbiota (Nugent 4-6; cyan) and BV-positive (Nugent 7-10; lavender). Pairwise comparisons between each infection group and the infection-negative control group were performed using Mann–Whitney U tests, and p-values are annotated above each violin. (B) Unadjusted forest plots showing β-coefficients (points) and 95% confidence intervals (whiskers) from single-predictor linear regression models assessing associations between each log-transformed cytokine and individual infections. Filled (black) circles indicate p < 0.05, and open (white) circles indicate p ≥ 0.05. (C) Adjusted forest plots showing multivariable linear regression models including all infections and vaginal microbiota categories simultaneously (CT, NG, TV, MG, *Candida* spp., intermediate microbiota, and BV-positive). β-coefficients therefore represent associations with each infection adjusted for co-infections. Filled (black) circles indicate p < 0.05, and open (white) circles indicate p ≥ 0.05. Abbreviations: BV, bacterial vaginosis. Alt Text: A three-panel figure showing distributions and regression analyses of cervicovaginal IL-1α, IL-1β, and IP-10. Panel A displays violin plots of cytokine concentrations across healthy controls and seven infection groups. Panels B and C show forest plots of unadjusted and adjusted beta-coefficients with 95% confidence intervals, highlighting that bacterial vaginosis is the strongest independent predictor of elevated interleukins and suppressed IP-10, while Candida spp. is independently associated with elevations in all three cytokines.

Conversely, IP-10 was significantly reduced in women with BV (median 33.5 vs. 146.3 pg/ml; p<0.0001) and intermediate microbiota (median 114.7 vs. 146.3 pg/ml; p=0.0463) compared to controls (Figure 1A). Meanwhile, *Candida* spp. (median 477.6 pg/ml; p<0.0001) and CT (median 190.6 pg/ml; p=0.0334) elevated IP-10, whereas TV, NG, and MG showed no association.

In multivariable analyses (adjusting for co-infections), BV, intermediate microbiota, *Candida* spp., and NG remained independently associated with elevated IL-1α and IL-1β (Figure 1C, Supplementary Table 1). CT was independently associated with elevated IL-1β (β = 0.38; 95% CI, 0.06–0.70; ∼2.4-fold increase) but not IL-1α. *Candida* spp., CT, and TV were independently associated with increased IP-10, whereas BV and intermediate microbiota independently predicted reduced IP-10 (Figure 1C).

### Population-level contribution of RTIs to genital inflammation

PAF analyses confirmed BV as the dominant population-level driver of elevated IL-1 (Figure 2, Supplementary Table 2). BV (Nugent 7–10) carried a 4.6-fold increased risk of elevated IL-1α (95% CI 2.8–7.8), accounting for 64.7% of high IL-1α responses; intermediate microbiota (Nugent 4–6) increased risk 2.6-fold (95% CI 1.4–4.8), contributing 23.5% (Figure 2A). For IL-1β, BV carried a 4.1-fold increased risk (95% CI 2.5–6.7), accounting for 60.5% of high responses, with intermediate microbiota contributing 23.8% (Figure 2B). Conversely, *Candida* spp. was the strongest predictor of elevated IP-10 (RR 2.2, 95% CI 1.5–3.4; PAF 7.8%), while BV predicted reduced IP-10 (RR 0.3, 95% CI 0.2–0.5), accounting for 51.4% of lower IP-10 responses (Figure 2C).

**Figure 2.**
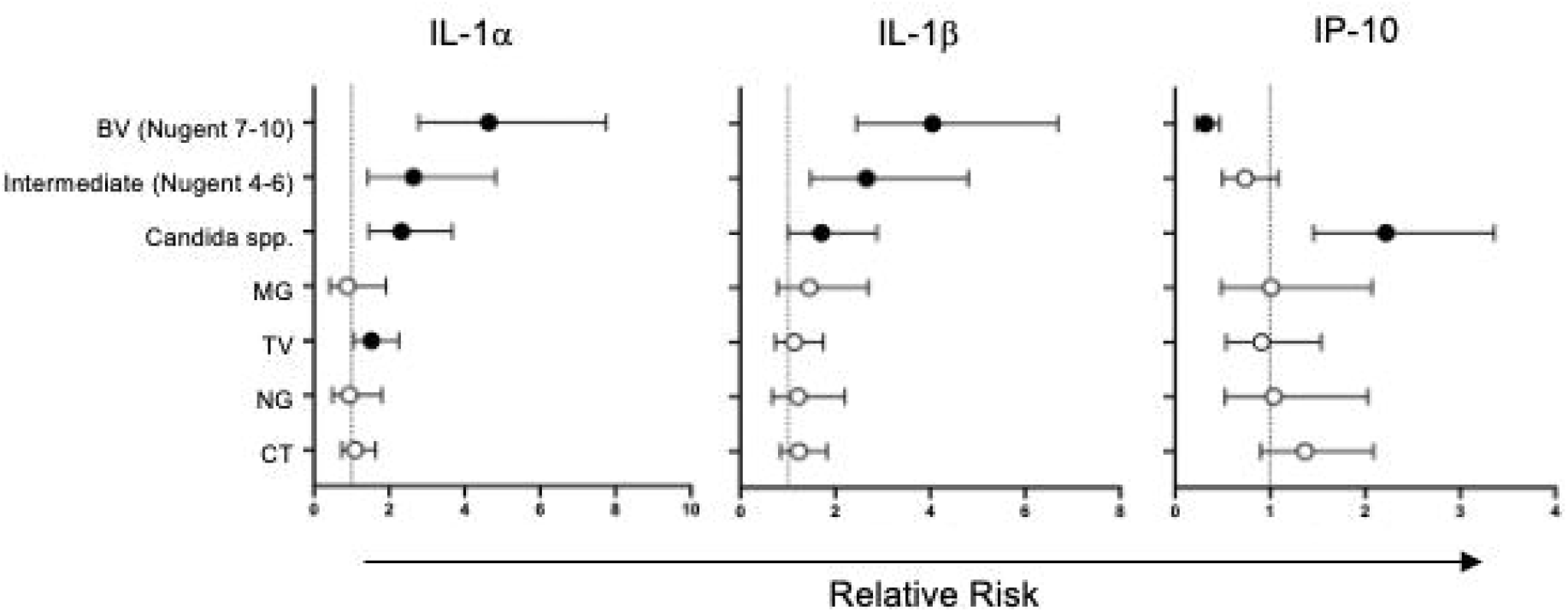
Forest plots of adjusted relative risks (RR) for elevated genital tract cytokine responses. Adjusted RRs and 95% confidence intervals for being in the top 25% (“high”) of log-transformed IL-1α (A), IL-1β (B), and IP-10 (C) distributions, associated with CT, NG, TV, MG, *Candida* spp., intermediate microbiota (Nugent 4–6), and Nugent-positive BV (7–10). RRs were estimated from multivariable Poisson regression models and are shown as circles with horizontal lines indicating 95% confidence intervals. Filled (black) circles indicate p < 0.05, and open (white) circles indicate p ≥ 0.05. The vertical dashed line at RR = 1 denotes no association; points to the right suggest increased risk of a high cytokine response, while those to the left suggest decreased risk. Corresponding population attributable fractions (PAFs) are presented in Supplementary Table 1. Alt Text: A three-panel forest plot showing adjusted relative risks (RRs) and 95% confidence intervals of having an elevated (top 25%) cytokine response for IL-1α (A), IL-1β (B), and IP-10 (C) associated with specific genital infections and vaginal dysbiosis. The plots highlight bacterial vaginosis as the strongest independent risk factor for elevated IL-1α and IL-1β, while Candida spp. is the strongest independent predictor of elevated IP-10.

### Impact of co-infections, Nugent-STI interactions, and HIV status on cytokine levels

Adjusting our multivariable regression models for HIV status showed that HIV infection was not independently associated with elevated IL-1α, IL-1β, or IP-10 concentrations (all p > 0.70), and its inclusion did not affect the association of other vaginal pathogens or conditions with genital inflammation (Supplementary Table 3). Furthermore, we found no statistically significant interaction or effect modification between Nugent score categories and STI status on any of the three cytokines (all p > 0.15; Supplementary Figure 4).

To address whether the suppressive effect of BV on IP-10 overrides the stimulatory effect of *Candida* spp., we compared cervicovaginal IP-10 concentrations in women with co-infections to those with single infections and healthy controls (Figure 3). Among STI-negative women, IP-10 levels in the co-infection group (median 200.6 pg/ml; IQR 34.6–350.4) were significantly higher than in women with BV only (median 33.5 pg/ml; IQR 5.8–96.7; p = 0.0049) and lower than in women with *Candida* only (median 477.6 pg/ml; IQR 178.9–649.4; p = 0.12). Notably, IP-10 concentrations in women with BV and *Candida* co-infection were statistically indistinguishable from those observed in healthy controls (median 146.3 pg/ml; IQR 85.9–244.2; p = 0.69), demonstrating that the opposing immunological effects of these two vaginal conditions counteract each other.

**Figure 3.**
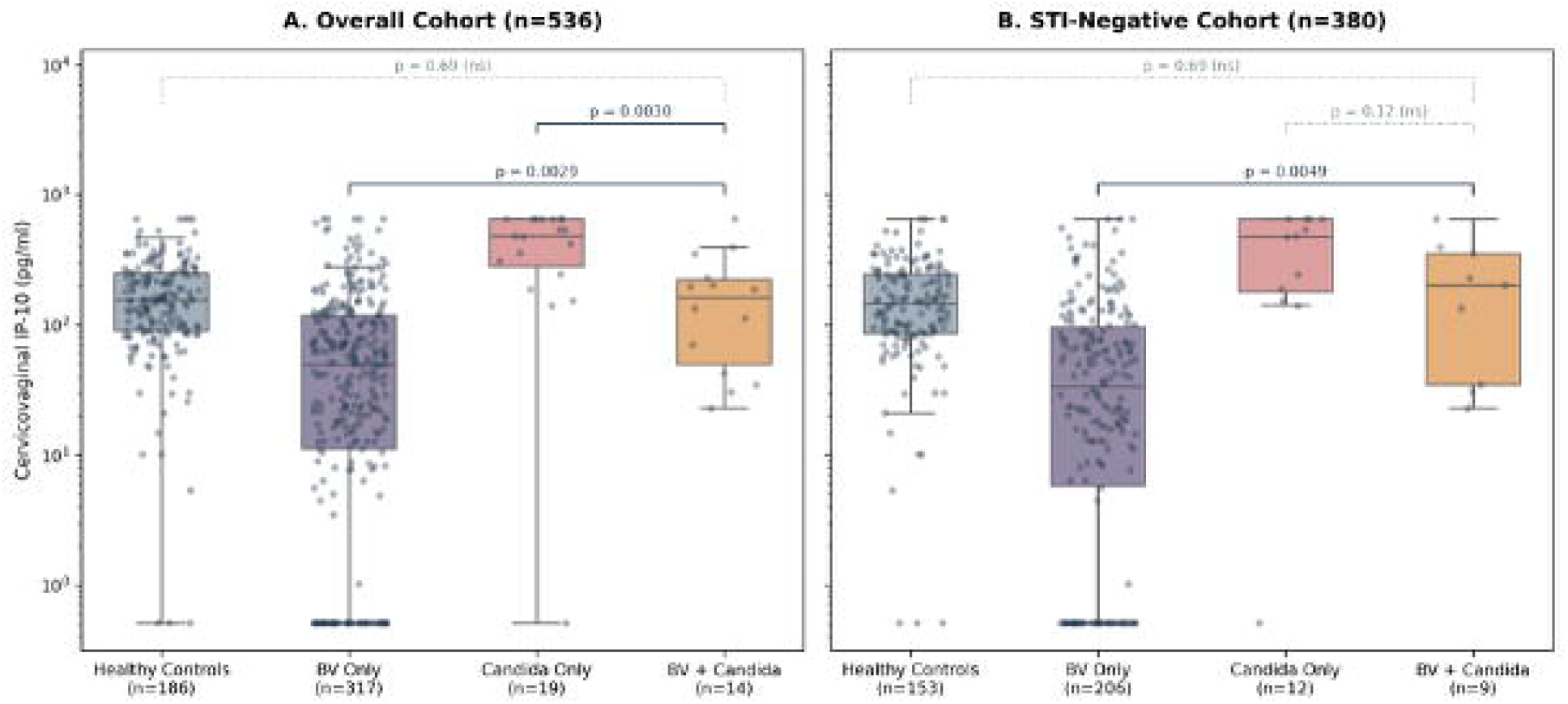
Cervicovaginal IP-10 concentrations in women with BV and *Candida* spp. co-infection. Boxplots show median IP-10 concentrations (with interquartile ranges and full data ranges) in STI-negative women stratified by BV status (Nugent score <4 vs. Nugent 7–10) and *Candida* detection by qPCR: Healthy controls (Nugent <4, *Candida*-negative; blue), BV only (Nugent 7–10, *Candida*-negative; purple), *Candida* only (Nugent <4, *Candida*-positive; pink), and co-infection (Nugent 7–10, *Candida*-positive; red). P-values from pairwise comparisons using Mann– Whitney U tests are shown. Abbreviations: BV, bacterial vaginosis. Alt Text: A boxplot comparing cervicovaginal IP-10 concentrations among STI-negative women across four groups: healthy controls (blue), BV-only (purple), Candida-only (pink), and BV-Candida co-infection (red). The plot illustrates that BV suppresses IP-10 and Candida elevates IP-10, while co-infected women show intermediate IP-10 levels that are statistically indistinguishable from healthy controls due to biological counter-regulation.

### Geographic variation in vaginal inflammation among RTI-negative women

Baseline cervicovaginal cytokines in RTI-negative women varied by country, being consistently lowest in South Africa and highest in Zimbabwe (Supplementary Figure 5A). IL-1α concentrations were lower in South Africa than Zimbabwe (p=0.0063), while IL-1β concentrations were lower in South Africa than Madagascar (p=0.0057) and Zimbabwe (p=0.0018). Similarly, IP-10 was lower in South Africa than Madagascar (p=0.0214) and Zimbabwe (p<0.0001).

### Geographic variation in RTI-associated responses

Across countries, CT, TV, BV, intermediate microbiota, and *Candida* spp. were consistently associated with elevated IL-1α and IL-1β compared to controls (Supplementary Figure 5B). NG and MG also elevated IL-1β, though IL-1α responses were less consistent. BV and intermediate microbiota predicted lower IP-10 across all sites. In contrast, *Candida* spp. was associated with increased IP-10 in South Africa and Zimbabwe, but not Madagascar, while TV-associated IP-10 reductions were only seen in Zimbabwe.

Although baseline cytokine levels and response magnitudes varied by country, infection-associated inflammatory patterns were conserved (Supplementary Figure 5B). Between-country comparisons showed minor variations in IL-1α responses to most RTIs and IL-1β responses to TV. Mixed-effects regression models (adjusting for confounders) confirmed that BV-, CT-, and TV-associated IL-1α responses were stronger in South Africa than Madagascar and Zimbabwe, as were IL-1β responses to TV (Supplementary Table 4). After adjustment, these country-level differences in cytokine responses persisted for most associations.

### Vaginal cytokine profiles by symptom status

Among RTI-negative women (n=154), 37.0% (57/154) reported genital symptoms (discharge or abdominal pain). Cytokine concentrations did not differ significantly between symptomatic and asymptomatic controls, with comparable IL-1α, IL-1β, and IP-10 in both groups (Figure 4). Among women with STIs only (n=33), 45.5% reported symptoms; cytokine concentrations did not differ by symptom status and were not elevated compared to asymptomatic controls (Figure 4A). Among women with BV only, both symptomatic (82/208) and asymptomatic (126/208) women had significantly higher IL-1α and IL-1β than controls (Figure 4B), while IP-10 was reduced in both groups. For women with *Candida* spp. only (n=12), IL-1α did not differ by symptom status or from controls. However, asymptomatic *Candida* was associated with elevated IL-1β and IP-10 (Figure 4C), despite the small sample size (6/12 asymptomatic).

**Figure 4.**
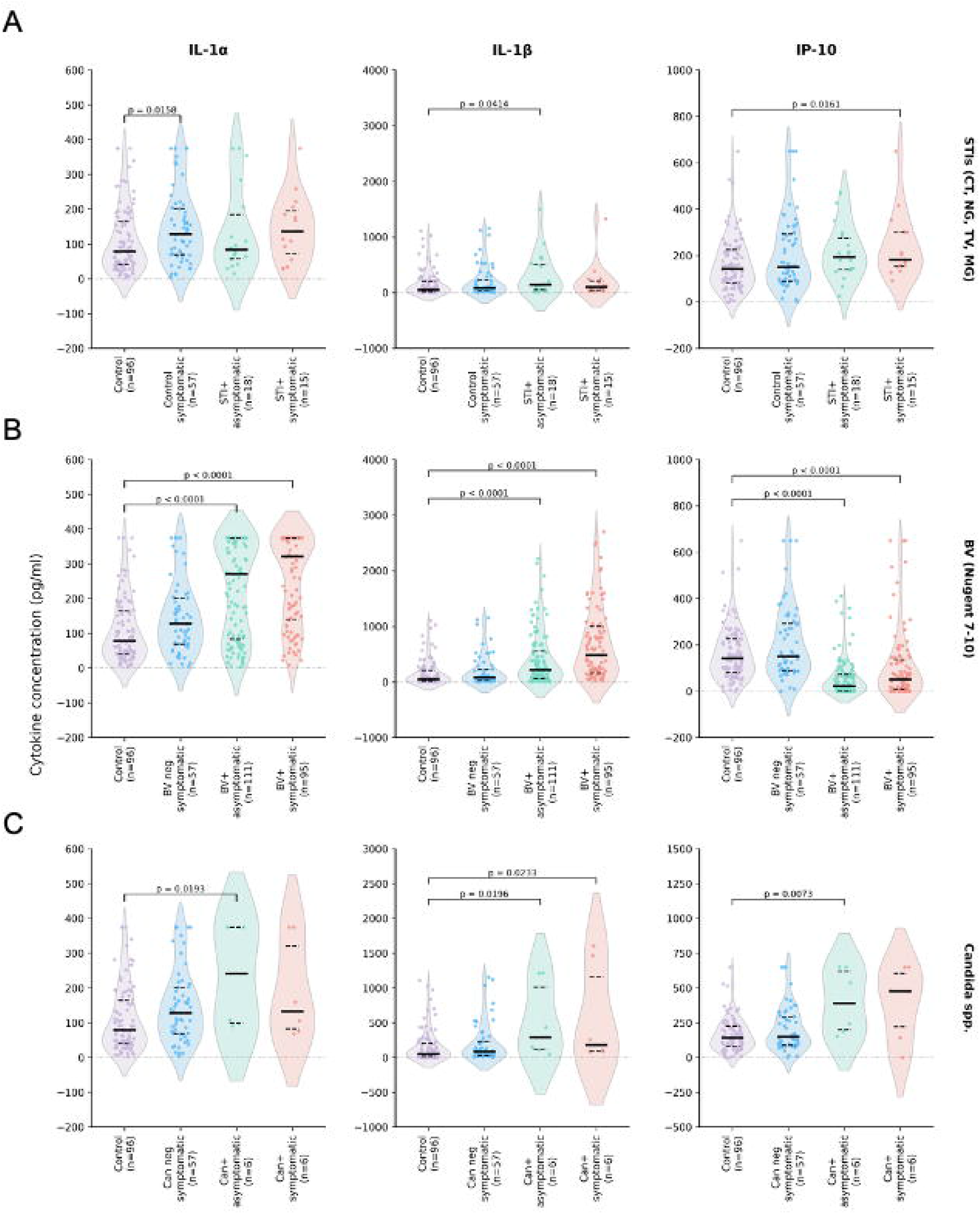
Cervicovaginal cytokine concentrations according to symptom status among women with STIs (A), BV (B), or *Candida* spp (C). (A) Cervicovaginal IL-1α, IL-1β, and IP-10 concentrations were compared between symptomatic and asymptomatic women with laboratory-confirmed STIs (n=33; Nugent score <4, *Candida*-negative). Asymptomatic, infection-negative controls (n=96) and symptomatic, infection-negative controls (n=57) are shown as references. (B) Cytokine concentrations <u>were</u> compared between symptomatic and asymptomatic women with BV only (n=206; STI-negative, *Candida*-negative). (C) Cytokine concentrations <u>were</u> compared between symptomatic and asymptomatic women with *Candida* spp. only (n=12; STI-negative, Nugent score <4). Each violin illustrates the full distribution of individual values, with the median (solid horizontal line) and interquartile range (dashed lines) overlaid. Comparisons between groups were performed using Mann–Whitney U tests. Abbreviations: BV, bacterial vaginosis; STI, sexually transmitted infection. Alt Text: A three-panel grid of violin plots comparing IL-1α, IL-1β, and IP-10 concentrations between healthy controls, symptomatic controls, and asymptomatic and symptomatic women with STIs (A), bacterial vaginosis (B), or Candida spp. (C). The plots illustrate that mucosal cytokine concentrations are highly elevated in infected women regardless of whether they report clinical symptoms, showing that inflammation is driven by infection status rather than the presence of symptoms.

## Discussion

In this multi-country study of women from South Africa, Madagascar and Zimbabwe, BV and intermediate vaginal microbiota were the dominant correlates of cervicovaginal inflammation, accounting for the largest proportion of elevated IL-1α and IL-1β responses at the population level. Women with intermediate microbiota exhibited inflammatory profiles similar to BV, suggesting that partial loss of Lactobacillus dominance has important biological consequences. Although RTIs exhibited pathogen-specific cytokine profiles, inflammatory signatures were broadly similar across countries and frequently occurred in the absence of symptoms.

These findings help contextualize the complex relationship between vaginal dysbiosis, genital inflammation, and HIV susceptibility. Elevated concentrations of IL-1α, IL-1β, and IP-10 are associated with increased HIV acquisition risk (2). In our study, women with dysbiosis generally had lower IP-10 concentrations despite substantial increases in IL-1α and IL-1β, suggesting that multiple inflammatory pathways independently contribute to mucosal vulnerability. Our finding that intermediate microbiota (Nugent 4–6) exhibits BV-like cytokine profiles suggests that mucosal inflammation arises before overt BV, indicating that current diagnostic frameworks underestimate the inflammatory consequences of partial Lactobacillus depletion (5, 29–31).

Symptom status was a poor indicator of mucosal inflammation (16); many women with substantial cytokine elevations were asymptomatic, while symptomatic women without RTIs showed no biomarker elevations. This highlights a fundamental limitation of syndromic management (1) and supports the potential of host-response screening to identify women with asymptomatic vaginal dysbiosis (32).

In multivariable diagnostic modeling, the three-biomarker panel combining IL-1α, IL-1β, and IP-10 provided a significantly better fit for detecting RTIs than any two-biomarker combination. Analysis of the individual contributions of these cytokines shows that while IL-1β is the primary driver of screening sensitivity for vaginal dysbiosis, the addition of IL-1α provides a critical gain in sensitivity by capturing active STIs that do not strongly induce IL-1β, without substantially increasing false-positive rates in healthy controls. This demonstrates that a multiplexed host-response screening approach can increase case detection (particularly for asymptomatic infections) while minimizing potential antibiotic overtreatment.

Several mechanisms may explain how dysbiosis-associated inflammation increases mucosal vulnerability, including epithelial barrier disruption and recruitment of activated HIV target cells (2, 4, 5, 20, 27, 28, 33, 34). Lactobacillus-deficient communities drive genital inflammation and target cell recruitment (5, 20, 33), whereas Lactobacillus dominance promotes immune quiescence (4, 5, 20, 27, 33). The dissociation between elevated IL-1 and lower IP-10 is consistent with studies showing that Lactobacillus casei produces lactocepin that degrades IP-10 (36), suggesting a similar suppressive mechanism by BV-associated bacteria.

Unlike most studies of genital inflammation, we evaluated Candida spp. and found independent associations with IL-1α, IL-1β, and IP-10. This aligns with evidence that asymptomatic Candida colonization promotes immune activation, including Th17-like cell recruitment and elevated inflammatory mediators (14). Given recent studies showing IL-1-driven immunopathology during vulvovaginal candidiasis (13, 37), Candida is an under-recognized contributor to immune activation that should be considered alongside other RTIs in mucosal health and diagnostic strategies.

Our finding that *Candida*-induced IP-10 elevation counteracts BV-induced IP-10 suppression in co-infected women has important clinical implications for host-response screening. In screening strategies that utilize IP-10 as a marker, co-infected women may present with ’normal’ IP-10 levels due to biological counter-regulation, potentially masking the infection. However, because both BV and *Candida* independently induce robust IL-1α and IL-1β secretion, the inclusion of these interleukin markers in a multiplexed screening panel ensures that active mucosal inflammation is still successfully detected. Such host-response signatures could support a multi-stage clinical triage workflow, where highly sensitive markers like IL-1α and IL-1β serve as an initial screen for active inflammation, while context-dependent markers like IP-10 guide differential management (distinguishing bacterial dysbiosis from targeted antimicrobial or antifungal needs).

The absence of a detectable cytokine signature for MG after adjustment for co-infections was notable. Although MG is recognised as a cause of cervicitis and upper reproductive tract disease (38–40), it was not associated with elevated cervicovaginal IL-1α, IL-1β, or IP-10 concentrations in multivariable analysis. However, MG was the least prevalent STI in this study, which may have limited statistical power to detect associations. This observation is consistent with previous studies suggesting that MG does not consistently induce strong genital tract inflammation and may instead establish persistent, low-grade or immune-modulating infections (38, 39, 41, 42). Further studies incorporating additional inflammatory and tissue-remodelling markers may be required to better characterise the host response to MG infection.

Baseline cytokine variability across countries may reflect differences in contraception, sexual activity, microbiome composition, or undetected pathogens (43–46). However, the consistency of infection-associated cytokine signatures despite this background variability supports their generalizability across diverse settings.

Limitations include a cross-sectional design and the omission of ulcerative STIs or Schistosoma haematobium, which is endemic in parts of the study region and causes female genital schistosomiasis (47). Additionally, a high proportion of women had IL-1α concentrations above the upper assay limit, potentially underestimating effect sizes. Future longitudinal studies should integrate cytokine profiling and microbiome sequencing to clarify host-microbe interactions.

In conclusion, vaginal dysbiosis was the dominant contributor to cervicovaginal inflammation in this multi-country cohort, with BV and intermediate microbiota driving the majority of the population-level inflammatory burden. Intermediate microbiota exhibited BV-like cytokine profiles, supporting the view that partial *Lactobacillus* depletion represents a biologically active dysbiotic state (19, 32). Despite background geographic variability and a lack of clinical symptoms in most cases, infection-associated signatures were consistent across countries. Collectively, these findings highlight the limitations of syndromic management and support host-response-based screening as a generalizable approach for identifying undetected RTIs and vaginal dysbiosis in African settings.

## Data Availability

All data produced in the present study are available upon reasonable request to the authors.

## Acknowledgements

We thank all women who participated in this study and the clinical, laboratory, and field teams in South Africa, Madagascar, and Zimbabwe. We acknowledge staff at collaborating institutions for recruitment, specimen collection, laboratory analyses, data management, and scientific input that supported study implementation and interpretation.

## GIFT study group and collaborators

Eneyi Kpokiri (London School of Hygiene & Tropical Medicine, United Kingdom), Bahiah Meyer (Division of Medical Virology, Department of Pathology, University of Cape Town, South Africa), Nangamso Cawe (Division of Medical Virology, Department of Pathology, University of Cape Town, South Africa), Phumla Radebe (Division of Medical Virology, Department of Pathology, University of Cape Town, South Africa), Conita Lombard (Division of Medical Virology, Department of Pathology, University of Cape Town, South Africa), Celia Mehou-Loko (Division of Medical Virology, Department of Pathology, University of Cape Town, South Africa), Yacoeb Ganief (Division of Medical Virology, Department of Pathology, University of Cape Town, South Africa), Rezeen Daniels (Desmond Tutu HIV Centre, University of Cape Town, South Africa), Anda Madikida (Desmond Tutu HIV Centre, University of Cape Town, South Africa), Mutsawashe Chisenga (Organization for Public Health Interventions and Development, Zimbabwe; The Biomedical Research and Training Institute, Zimbabwe), Joseph F Chipanga (Organization for Public Health Interventions and Development, Zimbabwe; The Biomedical Research and Training Institute, Zimbabwe), Tsitsi Bandason (Organization for Public Health Interventions and Development, Zimbabwe; The Biomedical Research and Training Institute, Zimbabwe), Patricia Makunyire (Organization for Public Health Interventions and Development, Zimbabwe; The Biomedical Research and Training Institute, Zimbabwe), Felicia Mhangami (Organization for Public Health Interventions and Development, Zimbabwe; The Biomedical Research and Training Institute, Zimbabwe), Jayjay Karumazondo (The Biomedical Research and Training Institute, Zimbabwe), Jason Naidoo (The Biomedical Research and Training Institute, Zimbabwe), Karen Webb (Organization for Public Health Interventions and Development, Zimbabwe), Lyndon Mungur (Medical Diagnostech, Cape Town, South Africa), Ashley Uys (Medical Diagnostech, Cape Town, South Africa), Darryl Uys (Medical Diagnostech, Cape Town, South Africa), Saberi Marias (Research Contracts and Innovation, University of Cape Town, South Africa), Solange Rasoanandrianina (Centre Hospitalier Universitaire de Gynécologie et Obstétrique de Befelatanana, Antananarivo, Madagascar), Dimitri Ravoavison (Centre Hospitalier Universitaire de Gynécologie et Obstétrique de Befelatanana, Antananarivo, Madagascar), Patrick Andry Rakotonirina (Centre Hospitalier Universitaire de Gynécologie et Obstétrique de Befelatanana, Antananarivo, Madagascar), Andrilaina Razakarivony (Centre Hospitalier Universitaire de Gynécologie et Obstétrique de Befelatanana, Antananarivo, Madagascar), Hantamalala Randria (Centre Hospitalier Universitaire de Gynécologie et Obstétrique de Befelatanana, Antananarivo, Madagascar), Alphonsine Rahantarimalala (Centre Hospitalier Universitaire de Gynécologie et Obstétrique de Befelatanana, Antananarivo, Madagascar), Lala Rafetrarivony (Institut Pasteur de Madagascar, Madagascar), Barivola Bernardson (Institut Pasteur de Madagascar, Madagascar), Tsiry Rasolofomanana (Institut Pasteur de Madagascar, Madagascar), Norohasina Randriamange (Institut Pasteur de Madagascar, Madagascar), Antsanirina Nomenjanahary (Institut Pasteur de Madagascar, Madagascar), Laurah Rabarisoa (Institut Pasteur de Madagascar, Madagascar), Dimitri Rasoloson (Institut Pasteur de Madagascar, Madagascar), Sahara Raveloson (Institut Pasteur de Madagascar, Madagascar), Laura Randrianantenaina (Institut Pasteur de Madagascar, Madagascar), Hanitratiavina Rakotonindriana (Institut Pasteur de Madagascar, Madagascar), Fenosoa Voanarivolalao (Institut Pasteur de Madagascar, Madagascar), Vaomalala Ranarinandra (Institut Pasteur de Madagascar, Madagascar), Farai Machinga (National Family Planning Council, Zimbabwe).

## Supplementary Figures

**Supplementary Figure 1**. Consort flow diagram of participant enrolment and data availability for the GIFT study.

Alt Text: A flowchart diagram illustrating the study design and participant flow. It outlines the numbers of women screened, enrolled, and ultimately analyzed for pathogens and cytokines across South Africa, Madagascar, and Zimbabwe, detailing the numbers and reasons for participant exclusion at each stage.

**Supplementary Figure 2**. Distribution of BV status and self-reported symptoms across countries. Alluvial plots illustrate the relationship between Nugent score category: no BV (0–3, teal), intermediate microbiota (4–6, orange), and BV-positive (7–10, purple) and the presence or absence of self-reported genital symptoms (yes/no) in South Africa (left), Madagascar (middle), and Zimbabwe (right). The width of each stream is proportional to the number of participants in that category.

Alt Text: Three side-by-side alluvial flow plots illustrating the relationship between Nugent score categories (no BV in teal, intermediate microbiota in orange, and BV-positive in purple) and self-reported genital symptoms (yes/no) for South Africa, Madagascar, and Zimbabwe. The width of the streams demonstrates that a large proportion of women with intermediate microbiota and bacterial vaginosis remain asymptomatic across all three settings.

**Supplementary Figure 3**. Pairwise co-infection frequencies among reproductive tract infections in the GIFT cohort. Values represent the number of participants with both infections. BV showed the highest degree of overlap with other infections, supporting adjustment for co-infections in multivariable analyses.

Alt Text: A triangular matrix grid showing the frequencies of pairwise co-infections between Chlamydia trachomatis, Neisseria gonorrhoeae, Trichomonas vaginalis, Mycoplasma genitalium, Candida spp., and bacterial vaginosis (BV). The matrix highlights that bacterial vaginosis is the most common co-infecting condition overlapping with other pathogens in the study population.

**Supplementary Figure 4**. Cervicovaginal cytokine concentrations stratified by Nugent category and STI status. Boxplots show log_10_-transformed concentrations of IL-1α (A), IL-1β (B), and IP-10 (C) in women stratified by Nugent category (normal Nugent 0–3, intermediate Nugent 4–6, BV-positive Nugent 7–10) and STI status (STI-negative, STI-positive). P-values for statistical interaction between Nugent categories and STI status are non-significant (p > 0.15).

Alt Text: A grid of boxplots showing log10-transformed concentrations of IL-1α (A), IL-1β (B), and IP-10 (C) in women stratified by Nugent category (normal Nugent 0–3, intermediate Nugent 4–6, BV-positive Nugent 7–10) and STI status (STI-negative versus STI-positive). The plots show parallel inflammatory trends across all categories, indicating no statistically significant interaction or effect modification between vaginal dysbiosis and STI status.

**Supplementary Figure 5**. Country-specific comparisons of IL-1α, IL-1β, and IP-10 concentrations in pathogen-positive and RTI-negative women. (A) Violin plots show cervicovaginal IL-1α, IL-1β, and IP-10 concentrations in RTI-negative controls (no laboratory-confirmed CT, NG, TV, or MG infection, no *Candida* detection, and Nugent score <4) from South Africa (blue), Madagascar (purple), and Zimbabwe (pink). Solid black lines indicate medians; dashed lines mark the 25th and 75th percentiles; whiskers denote the full data range. (B–E) Boxplots compare median cytokine concentrations (IL-1α, IL-1β, and IP-10) between RTI-negative (light shading) and pathogen-only-positive (dark shading) women within each country. Pathogen-positive groups were defined as women positive for the specified pathogen only, without co-infections or BV (Nugent <4). (F–H) Country-specific differences in IL-1α, IL-1β, and IP-10 concentrations for BV (Nugent 7–10), intermediate microbiota (Nugent 4–6), and *Candida* detection. Boxes show interquartile ranges, black lines mark medians, and whiskers indicate full ranges. Comparisons between groups were performed using Mann–Whitney U tests. P-values are shown only for statistically significant comparisons. Abbreviations: RTI, reproductive tract infection; BV, bacterial vaginosis.

Alt Text: A multi-panel figure comparing IL-1α, IL-1β, and IP-10 concentrations across South Africa, Madagascar, and Zimbabwe. Panel A displays violin plots of baseline cytokine levels in uninfected controls. Panels B through E display boxplots comparing uninfected controls and pathogen-only-positive women within each country, demonstrating that while baseline cytokine levels vary between countries, the inflammatory signatures associated with specific infections remain highly consistent across sites.

**Supplementary Figure 6**. Country-specific fold-change analysis of genital cytokine responses to RTIs, BV, and *Candida* colonisation. Boxplots show the fold-change in log-transformed median raw concentrations of IL-1α, IL-1β and IP-10, calculated as the ratio of median cytokine concentrations in pathogen-positive compared with pathogen-negative controls. Fold-change responses associated with: CT (top row left), NG (top row right), TV (second row left), MG (second row right), intermediate microbiota (Nugent scores 4–10; third row left), BV (Nugent scores 7– 10; third row right), and *Candida* colonisation (bottom row left). Analyses were performed separately in women from South Africa (blue), Madagascar (purple) and Zimbabwe (pink). Fold-change distributions were compared across countries using Kruskal–Wallis tests with Dunn’s multiple-comparison post hoc tests. Adjusted P-values <0.05 are shown. Boxes represent the interquartile range, horizontal lines indicate the median, and whiskers extend to the minimum and maximum values (range).

Alt Text: A grid of boxplots showing country-specific fold-changes of IL-1α, IL-1β, and IP-10 concentrations associated with Chlamydia trachomatis (CT), Neisseria gonorrhoeae (NG), Trichomonas vaginalis (TV), Mycoplasma genitalium (MG), intermediate microbiota, bacterial vaginosis (BV), and Candida colonisation. Within each panel, fold-change values relative to uninfected controls are shown separately for South Africa (blue), Madagascar (purple), and Zimbabwe (pink), demonstrating that the relative magnitude of cytokine responses to specific infections is highly consistent across geographic settings.

**Supplementary Table 1.**
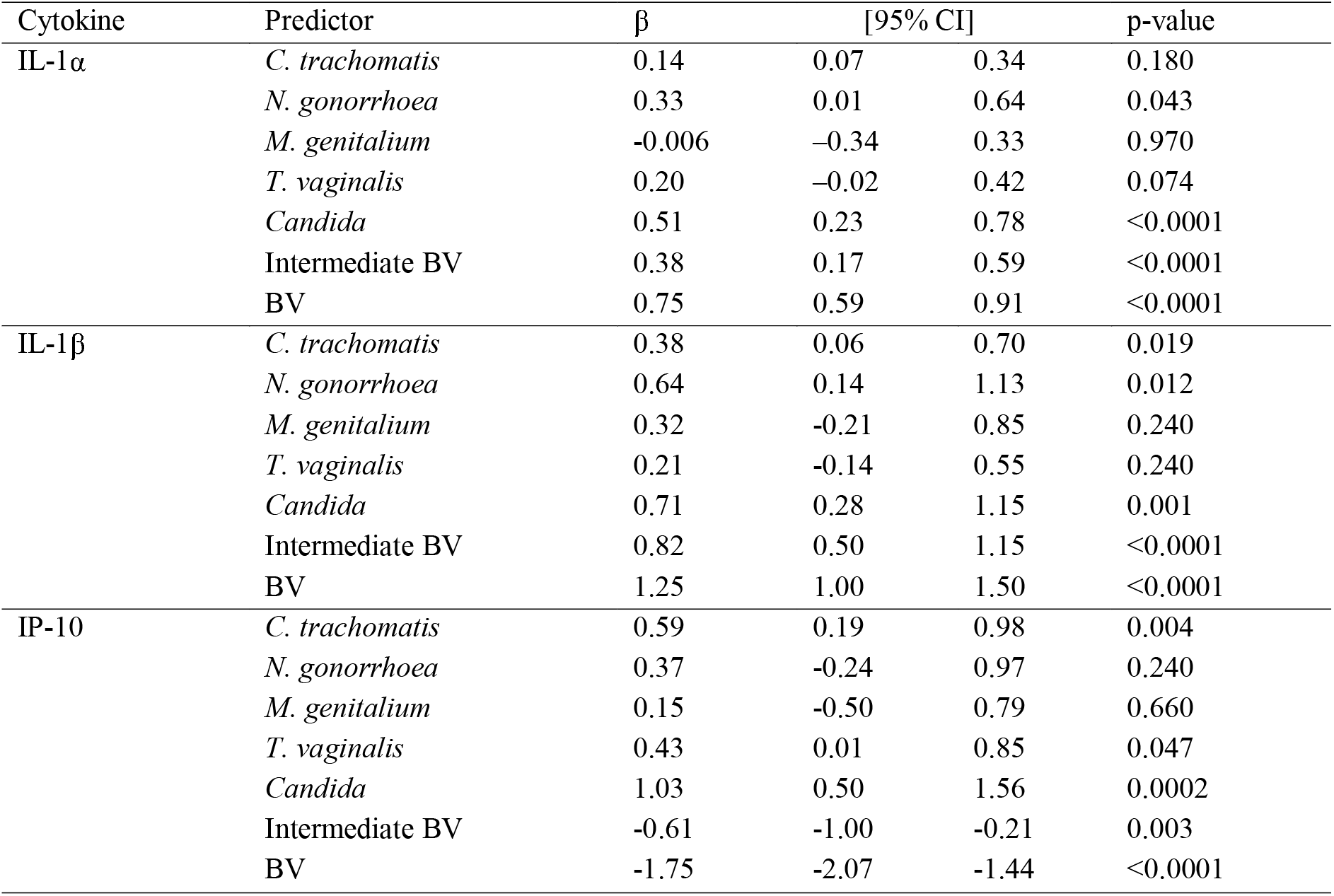
Associations between cytokines and vaginal tract infections, adjusting for co-infections.

**Supplementary Table 2.**
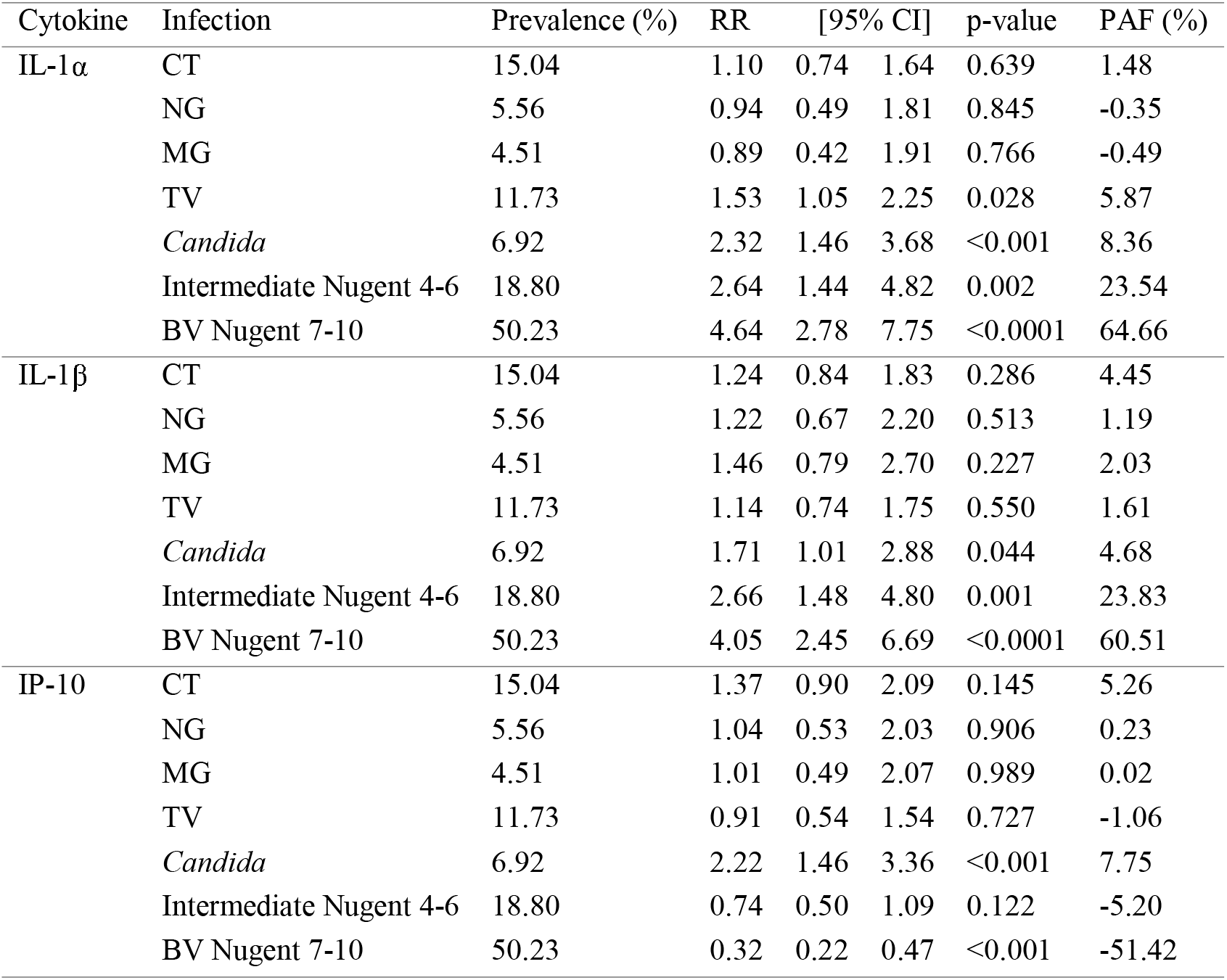
Adjusted relative risks (RRs) and population attributable fractions (PAFs) for elevated cervicovaginal cytokines.

**Supplementary Table 3:**
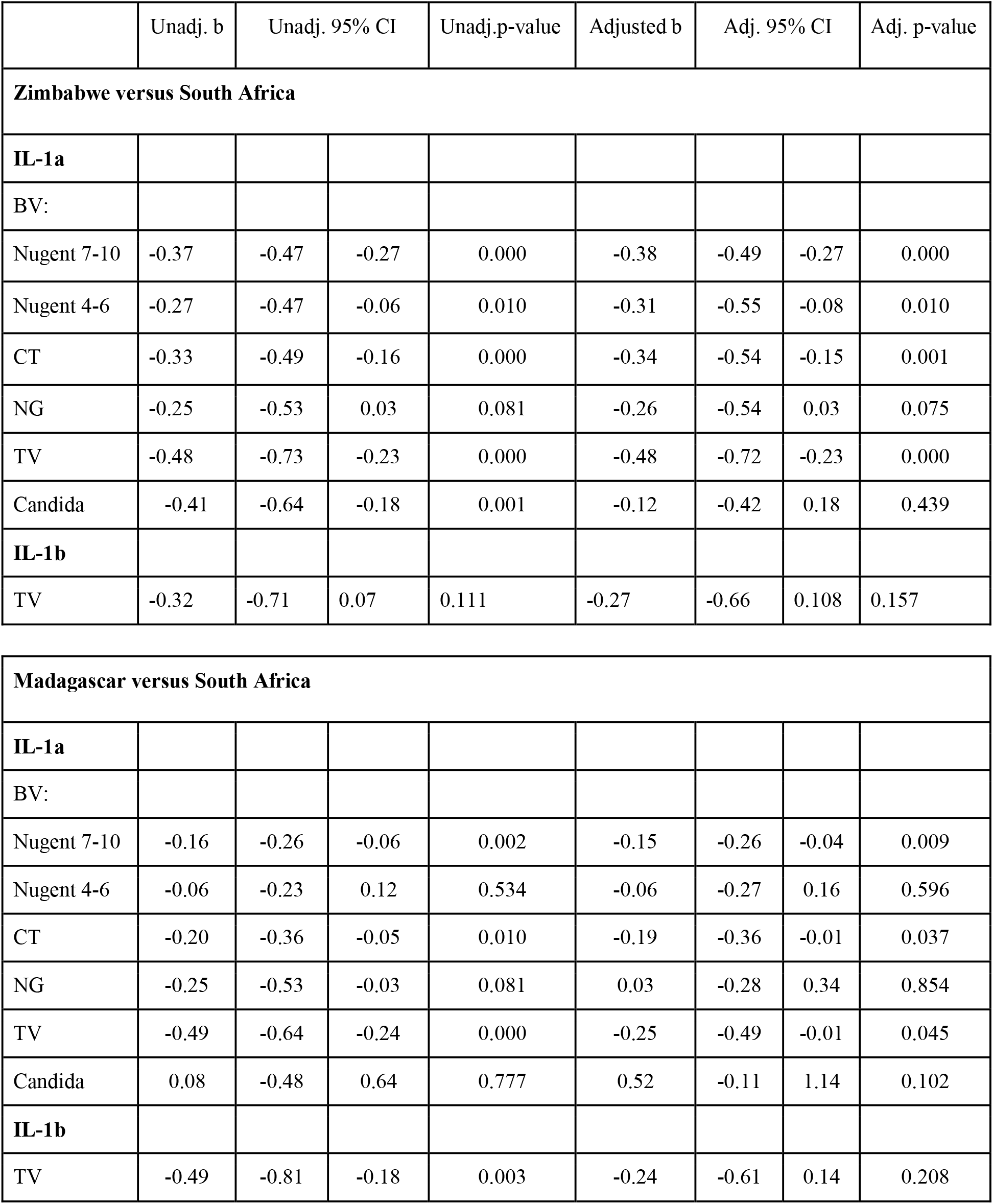
Mixed effects model estimates of IL-1α and IL-1β fold-change in Madagascar and Zimbabwe versus South Africa.

## References

1. Guidelines for the management of symptomatic sexually transmitted infections. WHO Guidelines Approved by the Guidelines Review Committee. Geneva 2021.

2. Masson L, Passmore JA, Liebenberg LJ, Werner L, Baxter C, Arnold KB, et al. Genital inflammation and the risk of HIV acquisition in women. Clin Infect Dis. 2015;61(2):260–9.

3. Arnold KB, Burgener A, Birse K, Romas L, Dunphy LJ, Shahabi K, et al. Increased levels of inflammatory cytokines in the female reproductive tract are associated with altered expression of proteases, mucosal barrier proteins, and an influx of HIV-susceptible target cells. Mucosal Immunol. 2016;9(1):194–205.

4. Masson L, Radzey N, Abrahams AG, Ngcapu S, McKinnon L, Jaspan HB. The vaginal microbiome and HIV acquisition risk. Lancet HIV. 2026;13(6):e414–e23.

5. Anahtar MN, Byrne EH, Doherty KE, Bowman BA, Yamamoto HS, Soumillon M, et al. Cervicovaginal bacteria are a major modulator of host inflammatory responses in the female genital tract. Immunity. 2015;42(5):965–76.

6. Michalow J, Hall L, Rowley J, Anderson RL, Hayre Q, Chico RM, et al. Prevalence of chlamydia, gonorrhoea, and trichomoniasis among male and female general populations in sub-Saharan Africa from 2000-2024: A systematic review and meta-regression analysis. medRxiv. 2024.

7. Schroder D, Sorano S, Shipitsyna E, Chaponda EB, Golparian D, Chikwanda E, et al. Prevalence and epidemiology of Mycoplasma genitalium and the absence of macrolide resistance in M. genitalium among pregnant women attending antenatal care in Zambia. Front Public Health. 2025;13:1576376.

8. Omosa-Manyonyi GS, Ponce IR, Rosati D, Bruno M, Kamau NW, Obimbo MM, et al. Genetic susceptibility to recurrent vulvovaginal candidiasis in an African population from Nairobi, Kenya. Sci Rep. 2025;15(1):12149.

9. Hillier SL. Diagnostic microbiology of bacterial vaginosis. Am J Obstet Gynecol. 1993;169(2 Pt 2):455–9.

10. Peebles K, Velloza J, Balkus JE, McClelland RS, Barnabas RV. High Global Burden and Costs of Bacterial Vaginosis: A Systematic Review and Meta-Analysis. Sex Transm Dis. 2019;46(5):304–11.

11. Sobel JD. Vulvovaginal candidosis. Lancet. 2007;369(9577):1961–71.

12. Yano J, Peters BM, Noverr MC, Fidel PL, Jr. Novel Mechanism behind the Immunopathogenesis of Vulvovaginal Candidiasis: “Neutrophil Anergy”. Infect Immun. 2018;86(3).

13. Cheng KO, Montano DE, Zelante T, Dietschmann A, Gresnigt MS. Inflammatory cytokine signalling in vulvovaginal candidiasis: a hot mess driving immunopathology. Oxf Open Immunol. 2024;5(1):iqae010.

14. Happel AU, Gasper M, Balle C, Konstantinus I, Gamieldien H, Dabee S, et al. Persistent, Asymptomatic Colonization with Candida is Associated with Elevated Frequencies of Highly Activated Cervical Th17-Like Cells and Related Cytokines in the Reproductive Tract of South African Adolescents. Microbiol Spectr. 2022;10(2):e0162621.

15. Zemouri C, Wi TE, Kiarie J, Seuc A, Mogasale V, Latif A, et al. The Performance of the Vaginal Discharge Syndromic Management in Treating Vaginal and Cervical Infection: A Systematic Review and Meta-Analysis. PLoS One. 2016;11(10):e0163365.

16. Mlisana K, Naicker N, Werner L, Roberts L, van Loggerenberg F, Baxter C, et al. Symptomatic vaginal discharge is a poor predictor of sexually transmitted infections and genital tract inflammation in high-risk women in South Africa. J Infect Dis. 2012;206(1):6–14.

17. Masson L, Barnabas S, Deese J, Lennard K, Dabee S, Gamieldien H, et al. Inflammatory cytokine biomarkers of asymptomatic sexually transmitted infections and vaginal dysbiosis: a multicentre validation study. Sex Transm Infect. 2019;95(1):5–12.

18. Jespers V, Kyongo J, Joseph S, Hardy L, Cools P, Crucitti T, et al. A longitudinal analysis of the vaginal microbiota and vaginal immune mediators in women from sub-Saharan Africa. Sci Rep. 2017;7(1):11974.

19. Masson L, Mlisana K, Little F, Werner L, Mkhize NN, Ronacher K, et al. Defining genital tract cytokine signatures of sexually transmitted infections and bacterial vaginosis in women at high risk of HIV infection: a cross-sectional study. Sex Transm Infect. 2014;90(8):580–7.

20. Srinivasan S, Richardson BA, Wallis JM, Fiedler TL, Strenk SM, Hoffman NG, et al. Vaginal Bacteria and Proinflammatory Host Immune Mediators as Biomarkers of Human Immunodeficiency Virus Acquisition Risk Among African Women. J Infect Dis. 2024;230(6):1444–55.

21. Gajer P, Brotman RM, Bai G, Sakamoto J, Schutte UM, Zhong X, et al. Temporal dynamics of the human vaginal microbiota. Sci Transl Med. 2012;4(132):132ra52.

22. Sun S, Wang H, Tsilimigras MC, Howard AG, Sha W, Zhang J, et al. Does geographical variation confound the relationship between host factors and the human gut microbiota: a population-based study in China. BMJ Open. 2020;10(11):e038163.

23. van Dorst M, Pyuza JJ, Nkurunungi G, Kullaya VI, Smits HH, Hogendoorn PCW, et al. Immunological factors linked to geographical variation in vaccine responses. Nat Rev Immunol. 2024;24(4):250–63.

24. Ramboarina S, Crucitti T, Gill K, Bekker LG, Harding-Esch EM, van de Wijgert J, et al. Novel point-of-care cytokine biomarker lateral flow test for the screening for sexually transmitted infections and bacterial vaginosis: study protocol of a multicentre multidisciplinary prospective observational clinical study to evaluate the performance and feasibility of the Genital InFlammation Test (GIFT). BMJ Open. 2024;14(5):e084918.

25. Mushi MF, Olum R, Bongomin F. Prevalence, antifungal susceptibility and etiology of vulvovaginal candidiasis in sub–Saharan Africa: a systematic review with meta-analysis and meta-regression. Medical Mycology. 2022; 60:myac037.

26. Jacot TA, Zalenskaya I, Mauck C, Archer DF, Doncel GF. TSPY4 is a novel sperm-specific biomarker of semen exposure in human cervicovaginal fluids; potential use in HIV prevention and contraception studies. Contraception. 2013;88(3):387–95.

27. Gosmann C, Anahtar MN, Handley SA, Farcasanu M, Abu-Ali G, Bowman BA, et al. Lactobacillus-Deficient Cervicovaginal Bacterial Communities Are Associated with Increased HIV Acquisition in Young South African Women. Immunity. 2017;46(1):29–37.

28. McClelland RS, Lingappa JR, Srinivasan S, Kinuthia J, John-Stewart GC, Jaoko W, et al. Evaluation of the association between the concentrations of key vaginal bacteria and the increased risk of HIV acquisition in African women from five cohorts: a nested case-control study. Lancet Infect Dis. 2018;18(5):554–64.

29. Munoz A, Hayward MR, Bloom SM, Rocafort M, Ngcapu S, Mafunda NA, et al. Modeling the temporal dynamics of cervicovaginal microbiota identifies targets that may promote reproductive health. Microbiome. 2021;9(1):163.

30. Joag V, Obila O, Gajer P, Scott MC, Dizzell S, Humphrys M, et al. Impact of Standard Bacterial Vaginosis Treatment on the Genital Microbiota, Immune Milieu, and Ex Vivo Human Immunodeficiency Virus Susceptibility. Clin Infect Dis. 2019;68(10):1675–83.

31. Manhanzva MT, Abrahams AG, Gamieldien H, Froissart R, Jaspan H, Jaumdally SZ, et al. Inflammatory and antimicrobial properties differ between vaginal Lactobacillus isolates from South African women with non-optimal versus optimal microbiota. Sci Rep. 2020;10(1):6196.

32. Muzny CA, Schwebke JR. Asymptomatic Bacterial Vaginosis: To Treat or Not to Treat? Curr Infect Dis Rep. 2020;22(12).

33. Klatt NR, Cheu R, Birse K, Zevin AS, Perner M, Noel-Romas L, et al. Vaginal bacteria modify HIV tenofovir microbicide efficacy in African women. Science. 2017;356(6341):938–45.

34. Lennard K, Dabee S, Barnabas SL, Havyarimana E, Blakney A, Jaumdally SZ, et al. Microbial Composition Predicts Genital Tract Inflammation and Persistent Bacterial Vaginosis in South African Adolescent Females. Infect Immun. 2018;86(1).

35. Kyongo JK, Crucitti T, Menten J, Hardy L, Cools P, Michiels J, et al. Cross-Sectional Analysis of Selected Genital Tract Immunological Markers and Molecular Vaginal Microbiota in Sub-Saharan African Women, with Relevance to HIV Risk and Prevention. Clin Vaccine Immunol. 2015;22(5):526–38.

36. Li Z, Levast B, Madrenas J. Staphylococcus aureus Downregulates IP-10 Production and Prevents Th1 Cell Recruitment. J Immunol. 2017;198(5):1865–74.

37. Coleman BM, Cook ME, Khan MR, Vogel AK, Wells AJ, Miao J, et al. An IL-1, IL-17, and IL-22 cytokine circuit controls vulvovaginal candidiasis independently of estrogen. PLoS Pathog. 2026;22(5):e1014202.

38. Lis R, Rowhani-Rahbar A, Manhart LE. Mycoplasma genitalium infection and female reproductive tract disease: a meta-analysis. Clin Infect Dis. 2015;61(3):418–26.

39. Taylor-Robinson D, Jensen JS. Mycoplasma genitalium: from Chrysalis to multicolored butterfly. Clin Microbiol Rev. 2011;24(3):498–514.

40. Manhart LE, Broad JM, Golden MR. Mycoplasma genitalium: should we treat and how? Clin Infect Dis. 2011;53 Suppl 3(Suppl 3):S129–42.

41. Qin L, Chen Y, You X. Subversion of the Immune Response by Human Pathogenic Mycoplasmas. Front Microbiol. 2019;10:1934.

42. Garza J, Gandhi K, Choi S, Sanchez A, Ventolini G. Cytokine profiles and Lactobacillus species presence in pre-menopausal subjects with genital Mycoplasma genitalium or Ureaplasma urealyticum colonization. Womens Health (Lond). 2021;17:17455065211009181.

43. Deese J, Masson L, Miller W, Cohen M, Morrison C, Wang M, et al. Injectable Progestin-Only Contraception is Associated With Increased Levels of Pro-Inflammatory Cytokines in the Female Genital Tract. Am J Reprod Immunol. 2015;74(4):357–67.

44. Herold BC, Mesquita PM, Madan RP, Keller MJ. Female genital tract secretions and semen impact the development of microbicides for the prevention of HIV and other sexually transmitted infections. Am J Reprod Immunol. 2011;65(3):325–33.

45. Boily-Larouche G, Lajoie J, Dufault B, Omollo K, Cheruiyot J, Njoki J, et al. Characterization of the Genital Mucosa Immune Profile to Distinguish Phases of the Menstrual Cycle: Implications for HIV Susceptibility. J Infect Dis. 2019;219(6):856–66.

46. Balle C, Konstantinus IN, Jaumdally SZ, Havyarimana E, Lennard K, Esra R, et al. Hormonal contraception alters vaginal microbiota and cytokines in South African adolescents in a randomized trial. Nat Commun. 2020;11(1):5578.

47. Rossi B, Previtali L, Salvi M, Gerami R, Tomasoni LR, Quiros-Roldan E. Female Genital Schistosomiasis: A Neglected among the Neglected Tropical Diseases. Microorganisms. 2024;12(3).

