## Supplemental Figures and Tables for "Distinct Cervicovaginal Cytokine Signatures Associated with Reproductive Tract Infections and Vaginal Dysbiosis Across Diverse Settings"

### Slide 1
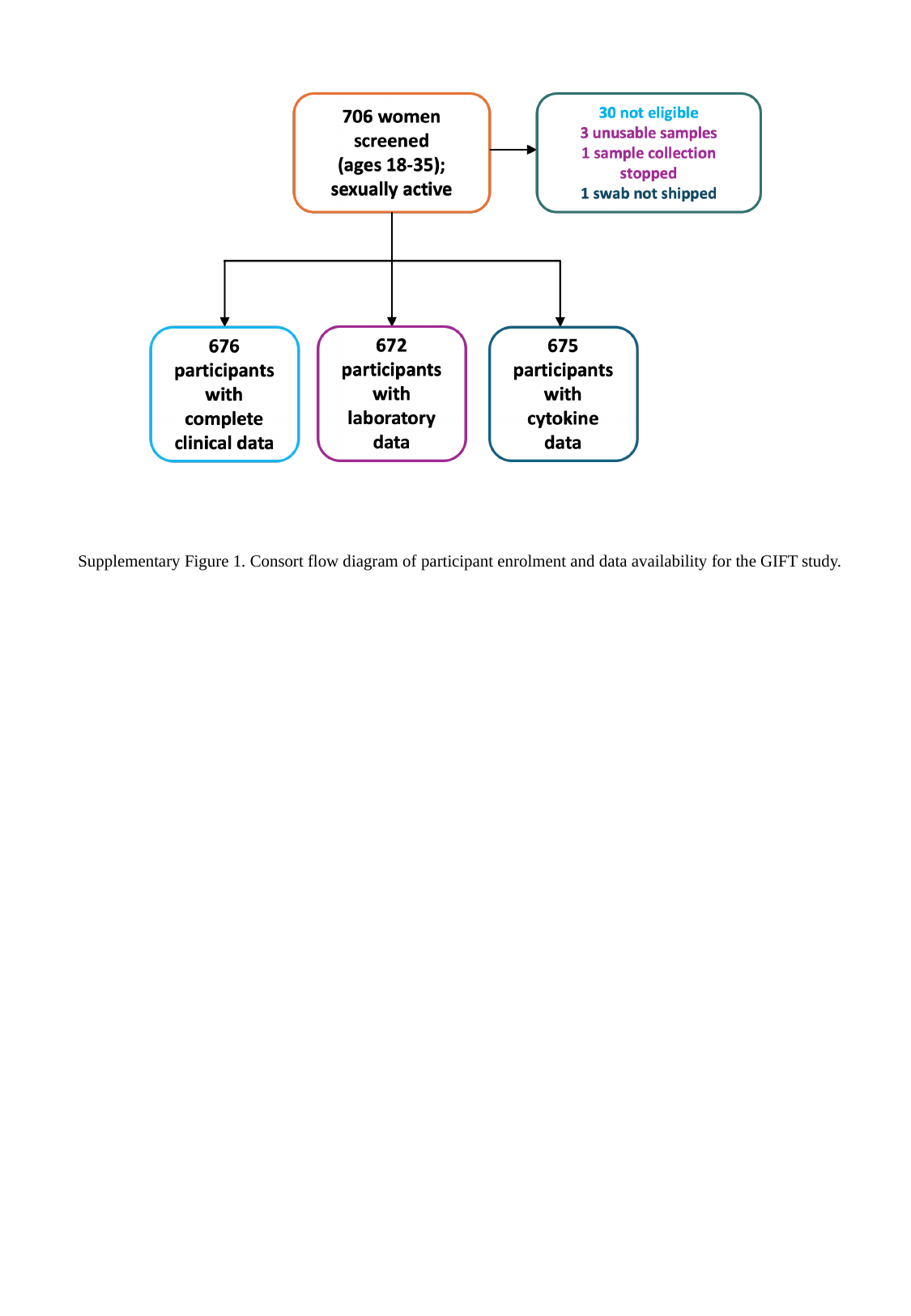

Supplementary Figure 1. Consort flow diagram of participant enrolment and data availability for the GIFT study.

### Slide 2
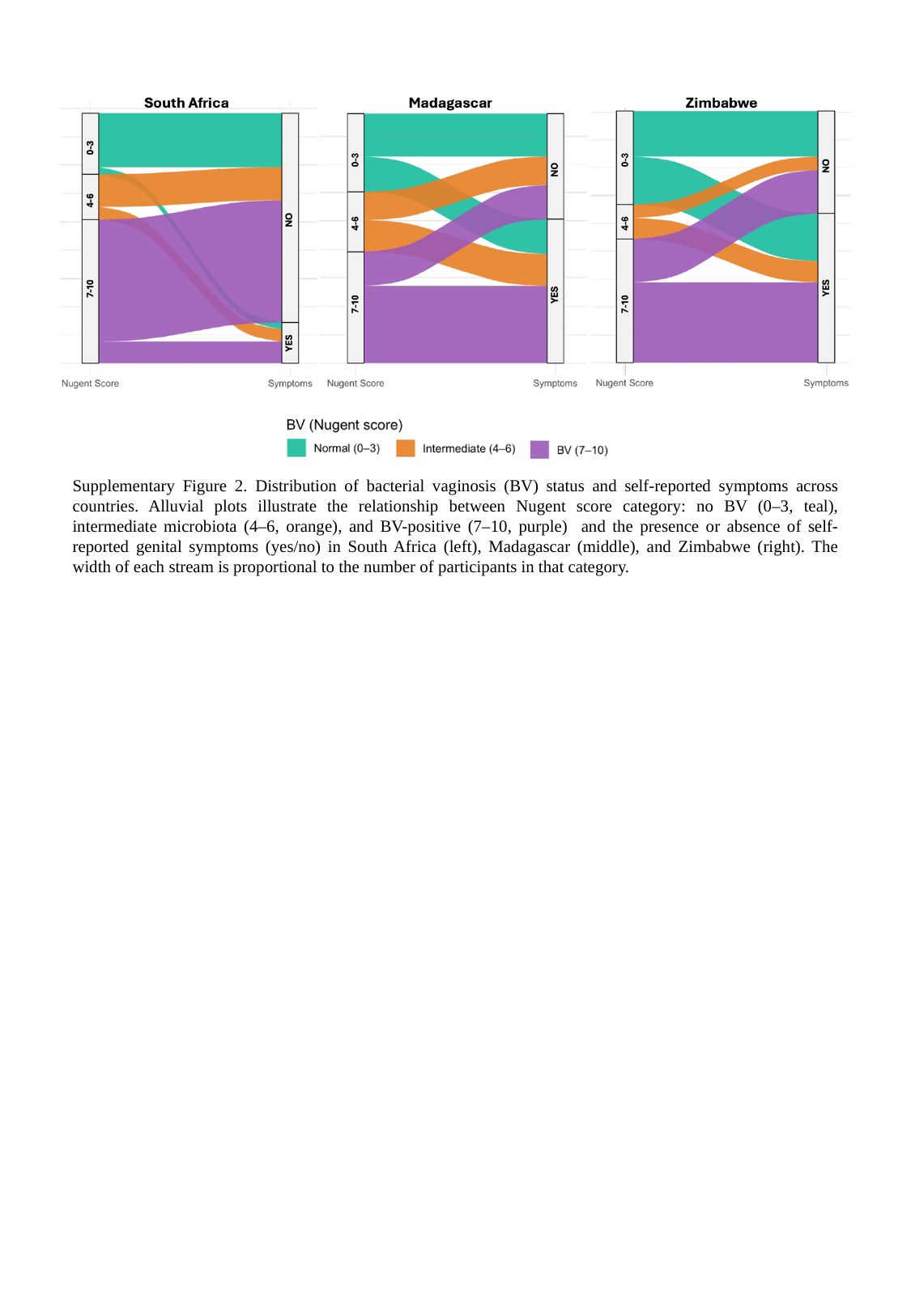

Supplementary Figure 2. Distribution of bacterial vaginosis (BV) status and self-reported symptoms across countries. Alluvial plots illustrate the relationship between Nugent score category: no BV (0–3, teal), intermediate microbiota (4–6, orange), and BV-positive (7–10, purple) and the presence or absence of self-reported genital symptoms (yes/no) in South Africa (left), Madagascar (middle), and Zimbabwe (right). The width of each stream is proportional to the number of participants in that category.

### Slide 3
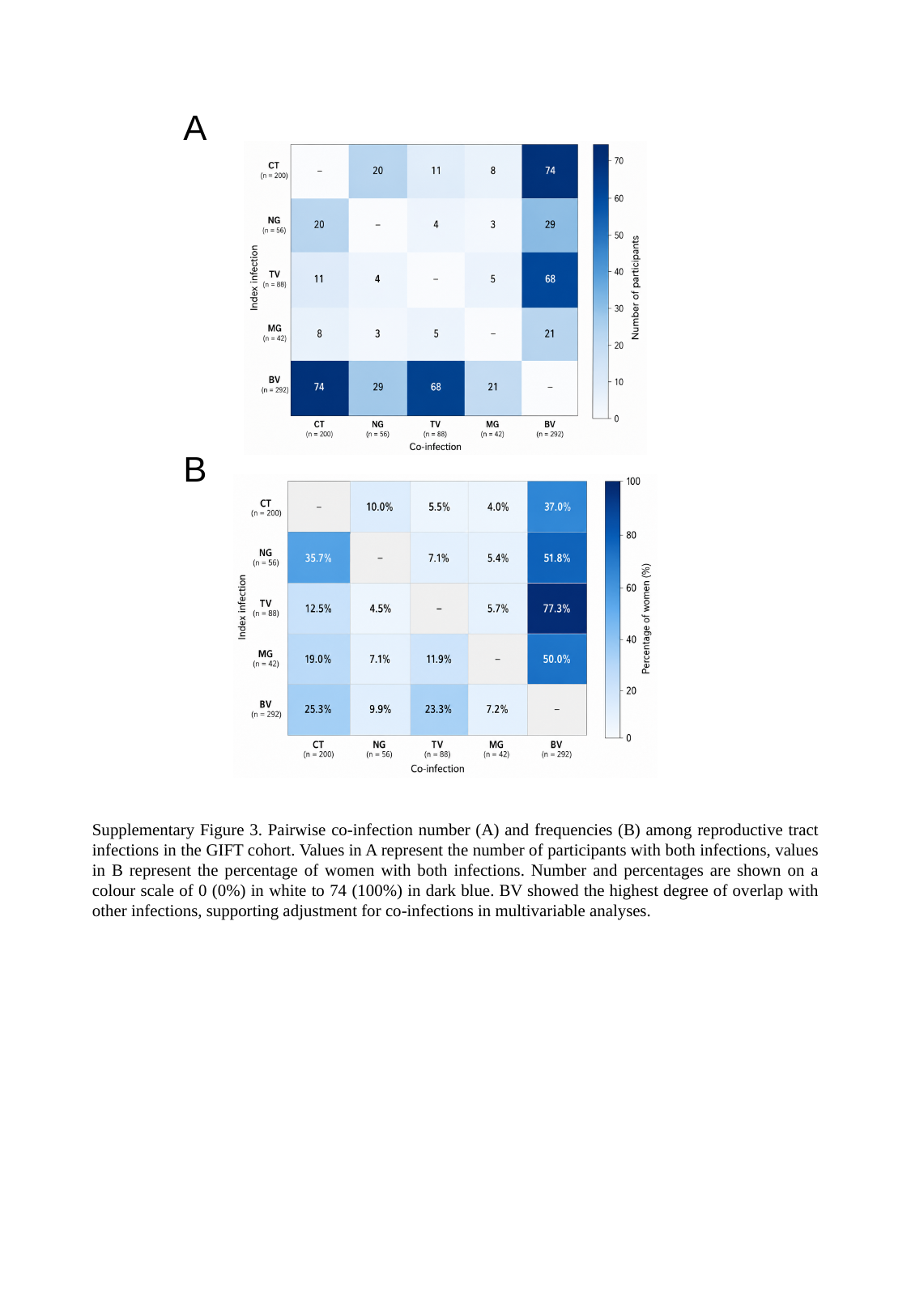

A
B
Supplementary Figure 3. Pairwise co-infection number (A) and frequencies (B) among reproductive tract infections in the GIFT cohort. Values in A represent the number of participants with both infections, values in B represent the percentage of women with both infections. Number and percentages are shown on a colour scale of 0 (0%) in white to 74 (100%) in dark blue. BV showed the highest degree of overlap with other infections, supporting adjustment for co-infections in multivariable analyses.

### Slide 4
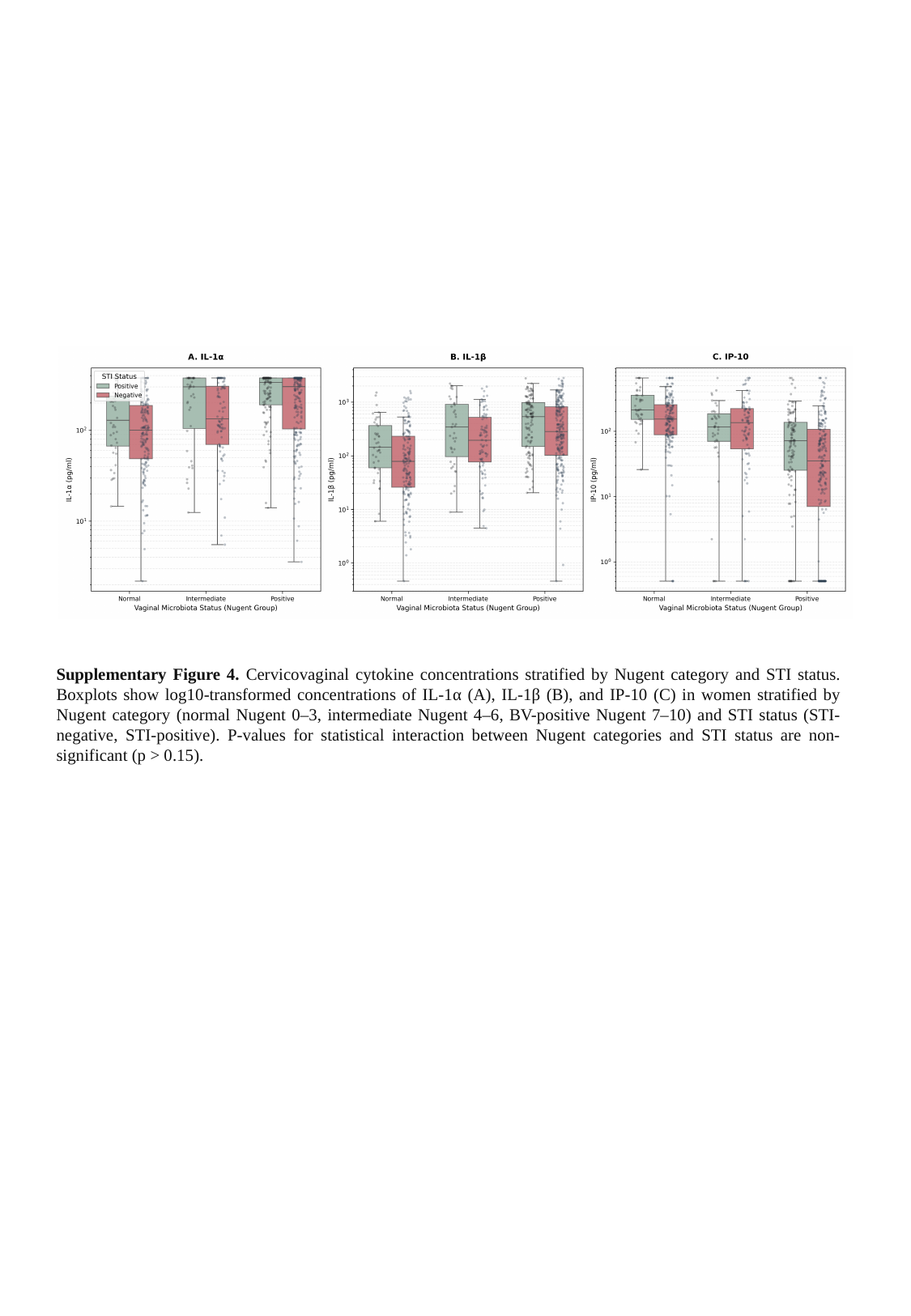

Supplementary Figure 4. Cervicovaginal cytokine concentrations stratified by Nugent category and STI status. Boxplots show log10-transformed concentrations of IL-1α (A), IL-1β (B), and IP-10 (C) in women stratified by Nugent category (normal Nugent 0–3, intermediate Nugent 4–6, BV-positive Nugent 7–10) and STI status (STI-negative, STI-positive). P-values for statistical interaction between Nugent categories and STI status are non-significant (p > 0.15).

### Slide 5
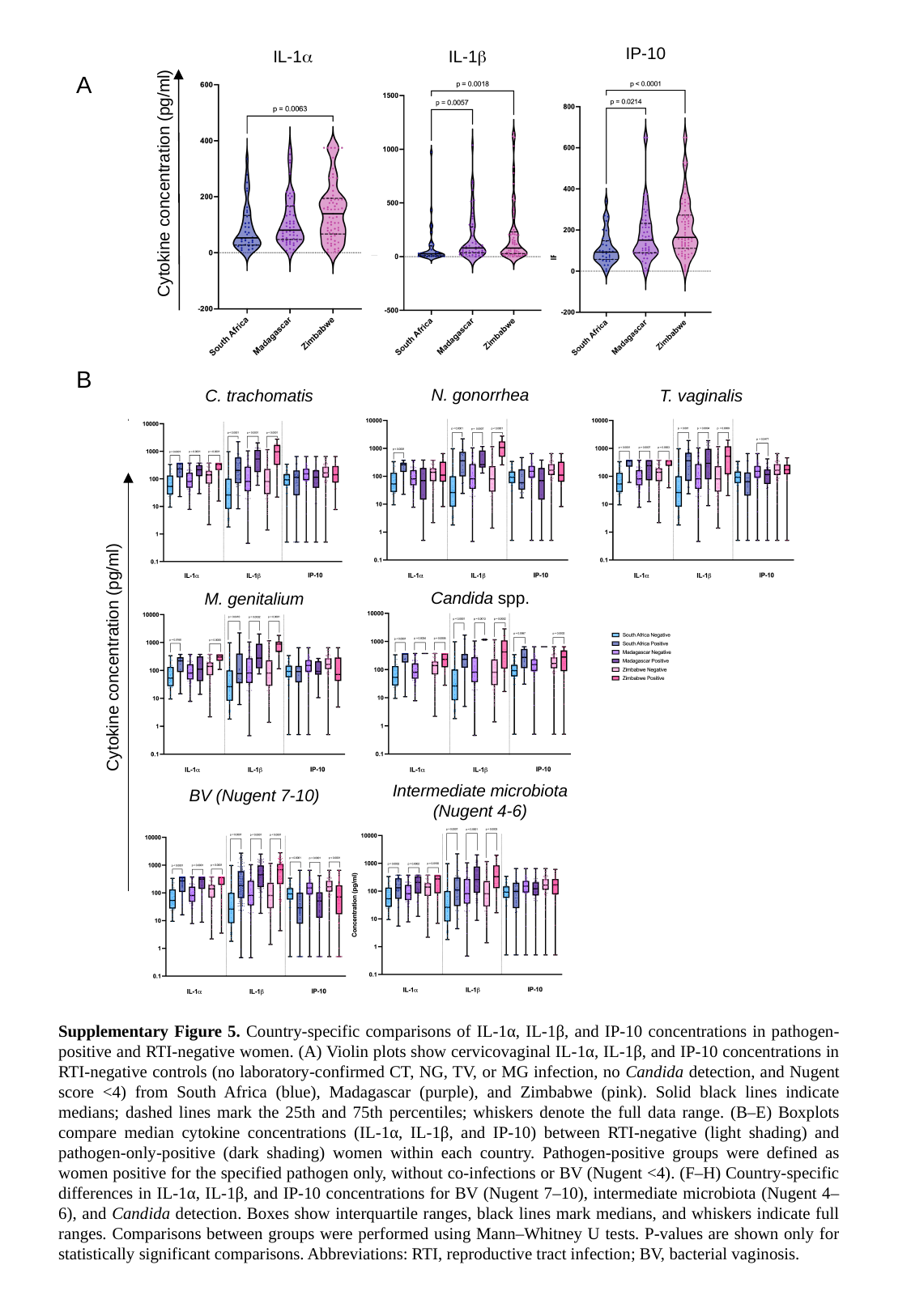

IP-10
IL-1α
IL-1β
A
Cytokine concentration (pg/ml)
B
N. gonorrhea
T. vaginalis
C. trachomatis
Candida spp.
M. genitalium
Cytokine concentration (pg/ml)
Intermediate microbiota (Nugent 4-6)
BV (Nugent 7-10)
Supplementary Figure 5. Country-specific comparisons of IL-1α, IL-1β, and IP-10 concentrations in pathogen-positive and RTI-negative women. (A) Violin plots show cervicovaginal IL-1α, IL-1β, and IP-10 concentrations in RTI-negative controls (no laboratory-confirmed CT, NG, TV, or MG infection, no Candida detection, and Nugent score <4) from South Africa (blue), Madagascar (purple), and Zimbabwe (pink). Solid black lines indicate medians; dashed lines mark the 25th and 75th percentiles; whiskers denote the full data range. (B–E) Boxplots compare median cytokine concentrations (IL-1α, IL-1β, and IP-10) between RTI-negative (light shading) and pathogen-only-positive (dark shading) women within each country. Pathogen-positive groups were defined as women positive for the specified pathogen only, without co-infections or BV (Nugent <4). (F–H) Country-specific differences in IL-1α, IL-1β, and IP-10 concentrations for BV (Nugent 7–10), intermediate microbiota (Nugent 4–6), and Candida detection. Boxes show interquartile ranges, black lines mark medians, and whiskers indicate full ranges. Comparisons between groups were performed using Mann–Whitney U tests. P-values are shown only for statistically significant comparisons. Abbreviations: RTI, reproductive tract infection; BV, bacterial vaginosis.

### Slide 6
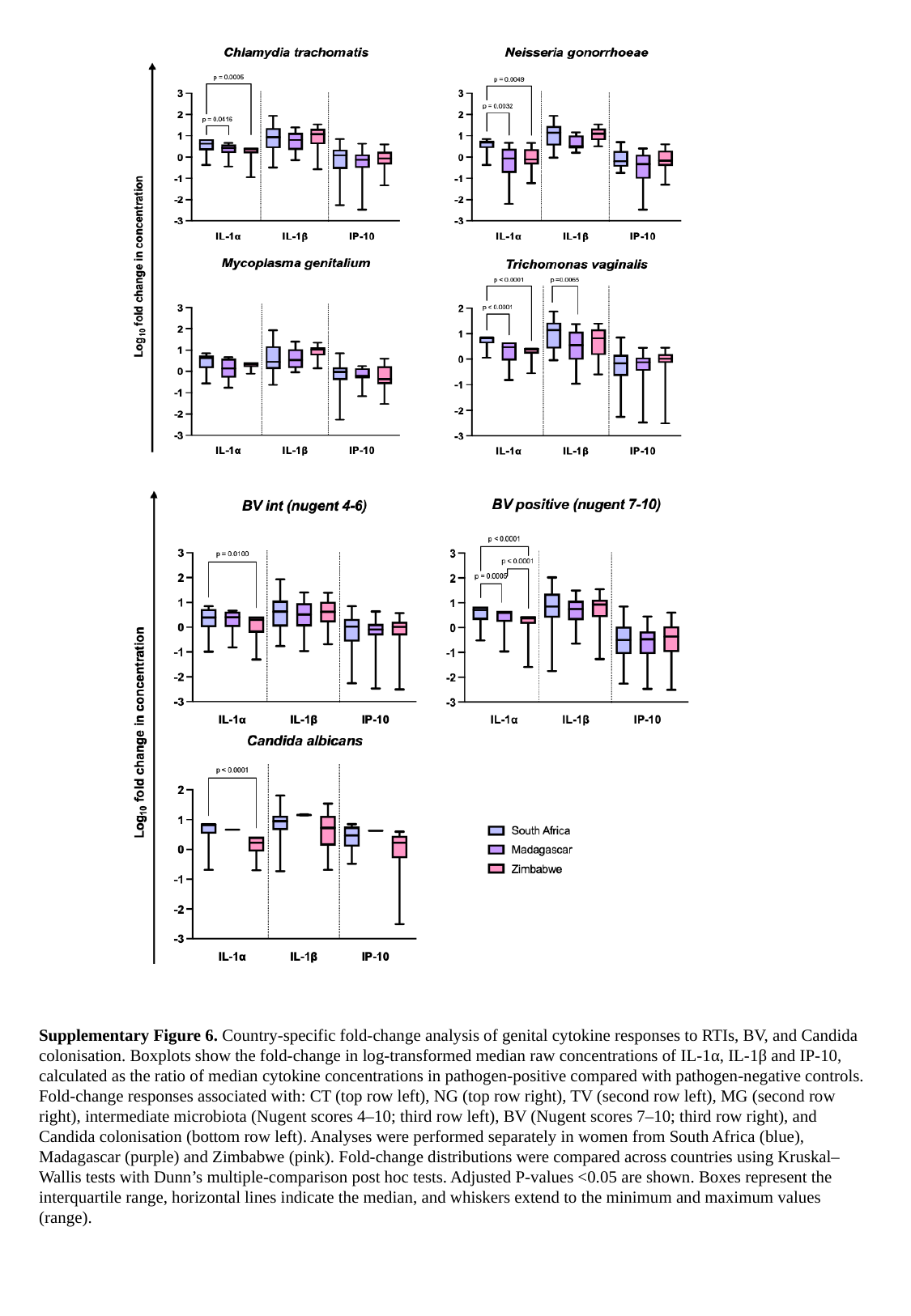

Supplementary Figure 6. Country-specific fold-change analysis of genital cytokine responses to RTIs, BV, and Candida colonisation. Boxplots show the fold-change in log-transformed median raw concentrations of IL-1α, IL-1β and IP-10, calculated as the ratio of median cytokine concentrations in pathogen-positive compared with pathogen-negative controls. Fold-change responses associated with: CT (top row left), NG (top row right), TV (second row left), MG (second row right), intermediate microbiota (Nugent scores 4–10; third row left), BV (Nugent scores 7–10; third row right), and Candida colonisation (bottom row left). Analyses were performed separately in women from South Africa (blue), Madagascar (purple) and Zimbabwe (pink). Fold-change distributions were compared across countries using Kruskal–Wallis tests with Dunn’s multiple-comparison post hoc tests. Adjusted P-values <0.05 are shown. Boxes represent the interquartile range, horizontal lines indicate the median, and whiskers extend to the minimum and maximum values (range).

### Slide 7
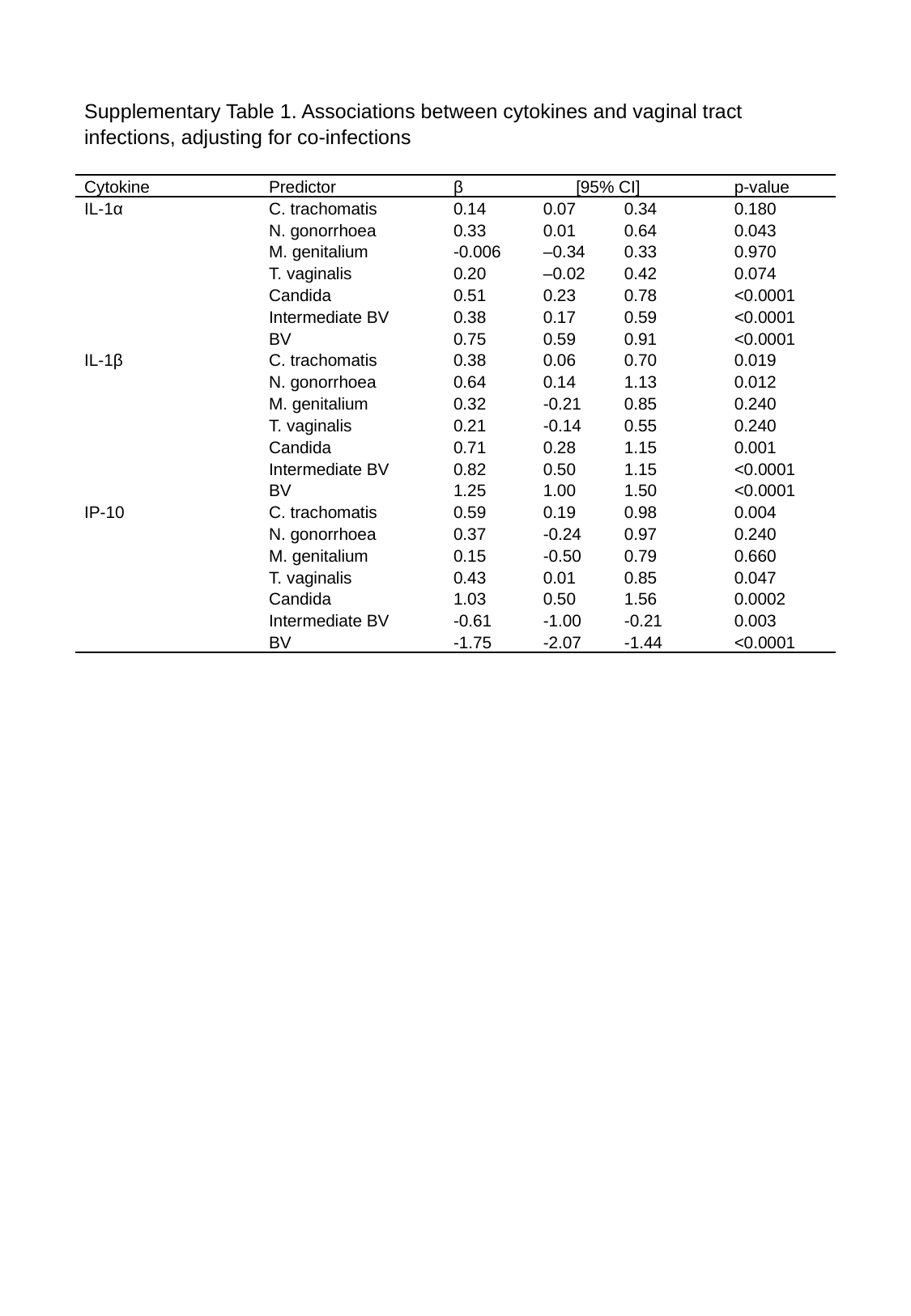

| Supplementary Table 1. Associations between cytokines and vaginal tract infections, adjusting for co-infections | | | | | |
| --- | --- | --- | --- | --- | --- |
| Cytokine | Predictor | β | [95% CI] | | p-value |
| IL-1α | C. trachomatis | 0.14 | 0.07 | 0.34 | 0.180 |
| | N. gonorrhoea | 0.33 | 0.01 | 0.64 | 0.043 |
| | M. genitalium | -0.006 | –0.34 | 0.33 | 0.970 |
| | T. vaginalis | 0.20 | –0.02 | 0.42 | 0.074 |
| | Candida | 0.51 | 0.23 | 0.78 | <0.0001 |
| | Intermediate BV | 0.38 | 0.17 | 0.59 | <0.0001 |
| | BV | 0.75 | 0.59 | 0.91 | <0.0001 |
| IL-1β | C. trachomatis | 0.38 | 0.06 | 0.70 | 0.019 |
| | N. gonorrhoea | 0.64 | 0.14 | 1.13 | 0.012 |
| | M. genitalium | 0.32 | -0.21 | 0.85 | 0.240 |
| | T. vaginalis | 0.21 | -0.14 | 0.55 | 0.240 |
| | Candida | 0.71 | 0.28 | 1.15 | 0.001 |
| | Intermediate BV | 0.82 | 0.50 | 1.15 | <0.0001 |
| | BV | 1.25 | 1.00 | 1.50 | <0.0001 |
| IP-10 | C. trachomatis | 0.59 | 0.19 | 0.98 | 0.004 |
| | N. gonorrhoea | 0.37 | -0.24 | 0.97 | 0.240 |
| | M. genitalium | 0.15 | -0.50 | 0.79 | 0.660 |
| | T. vaginalis | 0.43 | 0.01 | 0.85 | 0.047 |
| | Candida | 1.03 | 0.50 | 1.56 | 0.0002 |
| | Intermediate BV | -0.61 | -1.00 | -0.21 | 0.003 |
| | BV | -1.75 | -2.07 | -1.44 | <0.0001 |

### Slide 8
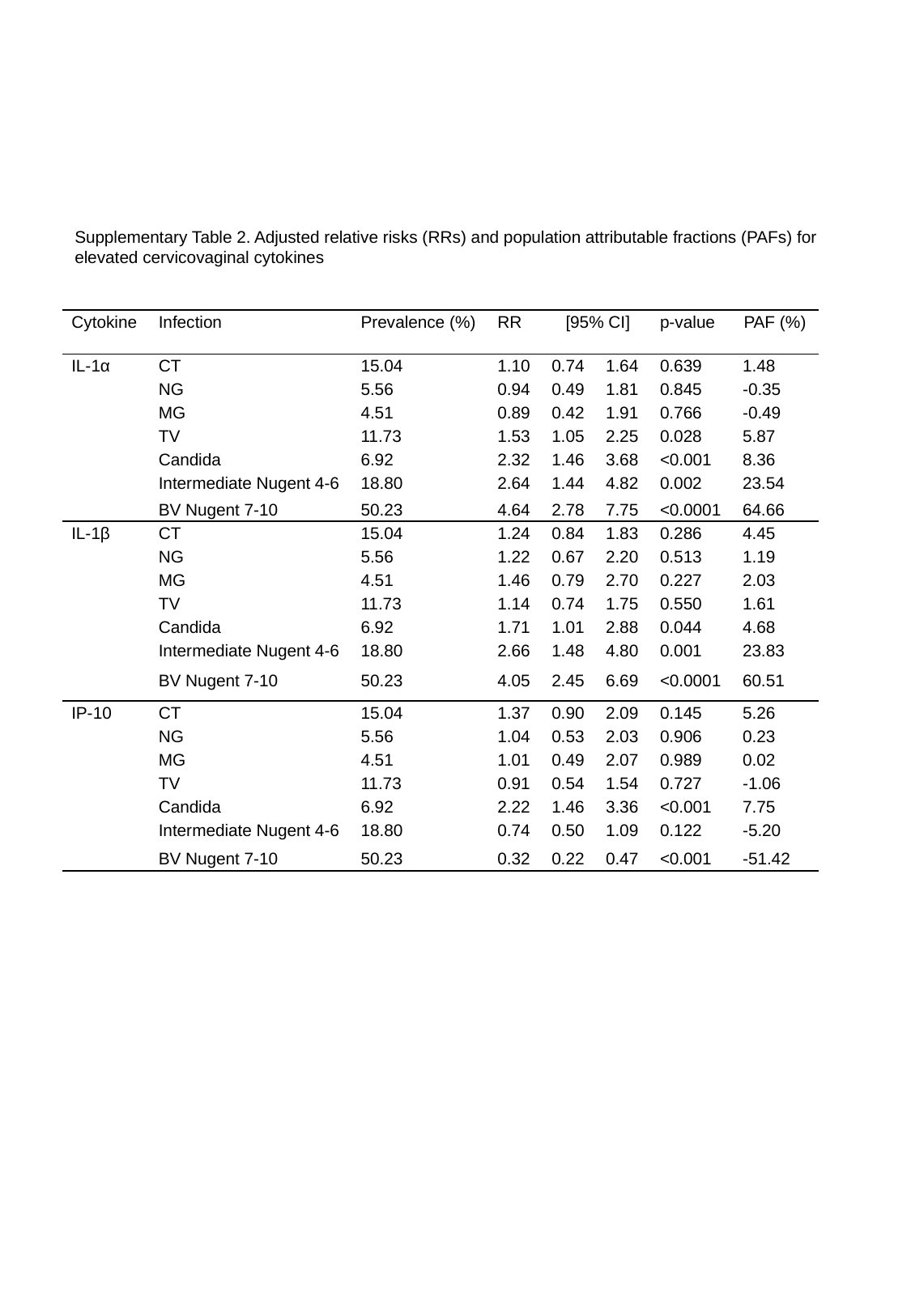

Supplementary Table 2. Adjusted relative risks (RRs) and population attributable fractions (PAFs) for elevated cervicovaginal cytokines
| Cytokine | Infection | Prevalence (%) | RR | [95% CI] | | p-value | PAF (%) |
| --- | --- | --- | --- | --- | --- | --- | --- |
| IL-1α | CT | 15.04 | 1.10 | 0.74 | 1.64 | 0.639 | 1.48 |
| | NG | 5.56 | 0.94 | 0.49 | 1.81 | 0.845 | -0.35 |
| | MG | 4.51 | 0.89 | 0.42 | 1.91 | 0.766 | -0.49 |
| | TV | 11.73 | 1.53 | 1.05 | 2.25 | 0.028 | 5.87 |
| | Candida | 6.92 | 2.32 | 1.46 | 3.68 | <0.001 | 8.36 |
| | Intermediate Nugent 4-6 | 18.80 | 2.64 | 1.44 | 4.82 | 0.002 | 23.54 |
| | BV Nugent 7-10 | 50.23 | 4.64 | 2.78 | 7.75 | <0.0001 | 64.66 |
| IL-1β | CT | 15.04 | 1.24 | 0.84 | 1.83 | 0.286 | 4.45 |
| | NG | 5.56 | 1.22 | 0.67 | 2.20 | 0.513 | 1.19 |
| | MG | 4.51 | 1.46 | 0.79 | 2.70 | 0.227 | 2.03 |
| | TV | 11.73 | 1.14 | 0.74 | 1.75 | 0.550 | 1.61 |
| | Candida | 6.92 | 1.71 | 1.01 | 2.88 | 0.044 | 4.68 |
| | Intermediate Nugent 4-6 | 18.80 | 2.66 | 1.48 | 4.80 | 0.001 | 23.83 |
| | BV Nugent 7-10 | 50.23 | 4.05 | 2.45 | 6.69 | <0.0001 | 60.51 |
| IP-10 | CT | 15.04 | 1.37 | 0.90 | 2.09 | 0.145 | 5.26 |
| | NG | 5.56 | 1.04 | 0.53 | 2.03 | 0.906 | 0.23 |
| | MG | 4.51 | 1.01 | 0.49 | 2.07 | 0.989 | 0.02 |
| | TV | 11.73 | 0.91 | 0.54 | 1.54 | 0.727 | -1.06 |
| | Candida | 6.92 | 2.22 | 1.46 | 3.36 | <0.001 | 7.75 |
| | Intermediate Nugent 4-6 | 18.80 | 0.74 | 0.50 | 1.09 | 0.122 | -5.20 |
| | BV Nugent 7-10 | 50.23 | 0.32 | 0.22 | 0.47 | <0.001 | -51.42 |

### Slide 9
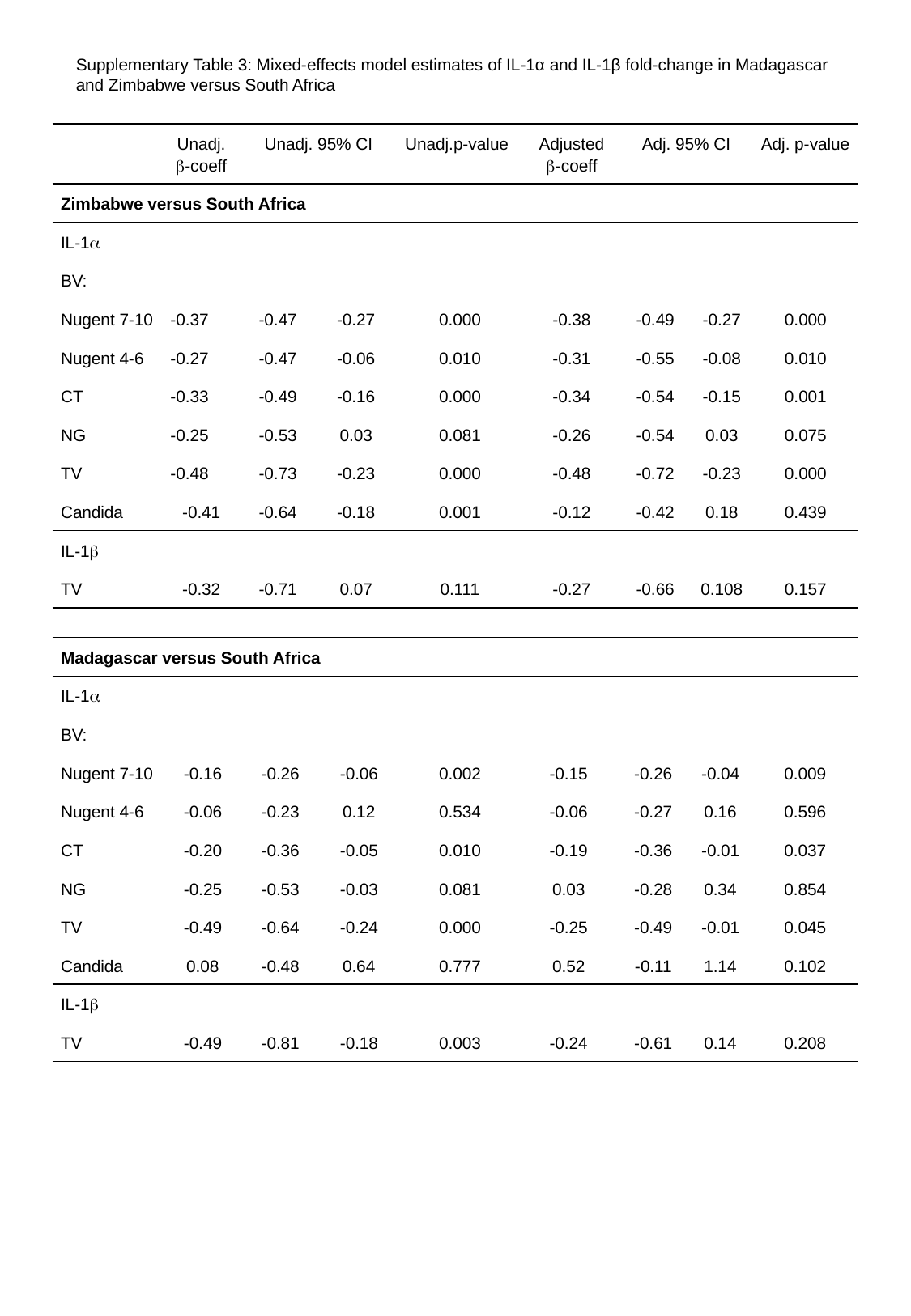

Supplementary Table 3: Mixed‐effects model estimates of IL-1α and IL-1β fold-change in Madagascar and Zimbabwe versus South Africa
| | Unadj. β-coeff | Unadj. 95% CI | | Unadj.p-value | Adjusted β-coeff | Adj. 95% CI | | Adj. p-value |
| --- | --- | --- | --- | --- | --- | --- | --- | --- |
| Zimbabwe versus South Africa | | | | | | | | |
| IL-1α | | | | | | | | |
| BV: | | | | | | | | |
| Nugent 7-10 | -0.37 | -0.47 | -0.27 | 0.000 | -0.38 | -0.49 | -0.27 | 0.000 |
| Nugent 4-6 | -0.27 | -0.47 | -0.06 | 0.010 | -0.31 | -0.55 | -0.08 | 0.010 |
| CT | -0.33 | -0.49 | -0.16 | 0.000 | -0.34 | -0.54 | -0.15 | 0.001 |
| NG | -0.25 | -0.53 | 0.03 | 0.081 | -0.26 | -0.54 | 0.03 | 0.075 |
| TV | -0.48 | -0.73 | -0.23 | 0.000 | -0.48 | -0.72 | -0.23 | 0.000 |
| Candida | -0.41 | -0.64 | -0.18 | 0.001 | -0.12 | -0.42 | 0.18 | 0.439 |
| IL-1β | | | | | | | | |
| TV | -0.32 | -0.71 | 0.07 | 0.111 | -0.27 | -0.66 | 0.108 | 0.157 |
| Madagascar versus South Africa | | | | | | | | |
| --- | --- | --- | --- | --- | --- | --- | --- | --- |
| IL-1α | | | | | | | | |
| BV: | | | | | | | | |
| Nugent 7-10 | -0.16 | -0.26 | -0.06 | 0.002 | -0.15 | -0.26 | -0.04 | 0.009 |
| Nugent 4-6 | -0.06 | -0.23 | 0.12 | 0.534 | -0.06 | -0.27 | 0.16 | 0.596 |
| CT | -0.20 | -0.36 | -0.05 | 0.010 | -0.19 | -0.36 | -0.01 | 0.037 |
| NG | -0.25 | -0.53 | -0.03 | 0.081 | 0.03 | -0.28 | 0.34 | 0.854 |
| TV | -0.49 | -0.64 | -0.24 | 0.000 | -0.25 | -0.49 | -0.01 | 0.045 |
| Candida | 0.08 | -0.48 | 0.64 | 0.777 | 0.52 | -0.11 | 1.14 | 0.102 |
| IL-1β | | | | | | | | |
| TV | -0.49 | -0.81 | -0.18 | 0.003 | -0.24 | -0.61 | 0.14 | 0.208 |
